# Conceptual framework of women’s food environments and determinants of food acquisition and dietary intakes in low- and middle-income countries: a systematic scoping review

**DOI:** 10.1101/2025.03.25.25324642

**Authors:** L O’Meara, J de Bruyn, T Hope, M Fajó-Pascual, R Hodge, C Turner, M Stoynova, K Wellard, E Ferguson, P Dominguez-Salas

## Abstract

**Background:** Progress on maternal health in low- and middle-income countries (LMICs) has stagnated, underscoring calls for holistic approaches to improve women’s nutrition. Diets link human health and environmental sustainability, necessitating equitable food system transformations to address climate change and malnutrition in all its forms. Food environments are a key entry point within food systems for improving nutrition; however, existing frameworks are not gender-sensitive and few consider vulnerable groups in low-resource settings.

**Methods:** We conducted a systematic scoping review of peer-reviewed literature published in English, Spanish, Portuguese, and French from Web of Science, EBSCO, and PubMed to identify determinants of food acquisition practices and dietary intakes of women of reproductive age in LMICs. We synthesised 518 studies from 125 countries. By systematically identifying 143 eco-social, structural and individual-level determinants, we identified key determinants to develop an empirically grounded food environment conceptual framework for women.

**Findings:** We identified women’s agency, characterised by decision-making and financial autonomy, bargaining power, control over time, and freedom of movement, as a prominent mediator of women’s food acquisition and dietary intakes, including across regions and the rural-urban continuum. Findings indicate that women’s agency, alongside supportive legislative, structural, and social enabling environments will be critical leverage points for improving women’s access to and consumption of nutritious foods, especially where resources are constrained.

**Conclusion:** For countries to sustainably address malnutrition, this empirically grounded framework identifies pathways for addressing the macro, social and individual determinants of food acquisition and dietary intakes that are often overlooked yet critical for resilient food environments and sustainable development.

**Key messages:**

- This is the first study to provide an empirically grounded food environment conceptual framework specific to women in LMICs.
- By systematically mapping key determinants of women’s food acquisition and dietary intakes, we identified novel food environment dimensions to develop an empirically grounded framework for women with applicability across regions and the rural-urban continuum in low- and middle-income countries.
- This novel conceptual framework, emphasising the importance of women’s agency in relation to external and personal food environments, may be used to guide research needs, analyses, and entry points for interventions to sustainably improve women’s nutrition in LMICs, especially in resource-constrained settings.

**Research in context:** *Evidence before this study:* Women in low- and middle-income countries (LMICs) are disproportionally affected by malnutrition in all its forms. Food environments are a key food system entry point to improve nutrition. However, current food environment frameworks are not gender-sensitive, limiting the effectiveness of nutrition interventions. We systematically searched Web of Science Core Collection, EBSCO, and PubMed for peer-reviewed studies published between 1^st^ January 2010 and 30^th^ April 2023 in English, Spanish, Portuguese, and French reporting on determinants of women’s food acquisition practices and dietary intakes in LMICs. We used a broad search criterion based on emerging food environment concepts and the expanded food security definition. Data were extracted for women aged 15-49 years and by physiological status (pregnant, lactating, and non-pregnant/non-lactating).

*Added value of this study:* This is the first study to provide an empirically grounded food environment conceptual framework specific to women in LMICs. By mapping patterns of 143 determinants inductively derived from the literature against existing conceptual frameworks, we identified novel determinants of women’s food acquisition and dietary intakes. Our results are representative across multiple geographical regions and the rural-urban continuum in LMICs. We add a novel socio-ecological layer: women’s agency as a key mediator for the ability of an individual to exert control over resources contributing to one’s own dietary outcomes.

*Implications of all the available evidence:* Women’s food environments are complex, necessitating holistic systems approaches to sustainably improve women’s nutrition in LMICs. It is critical that policies and programmes address underlying legislative, structural and socio-cultural determinants mediating women’s agency, alongside other key external and personal food environment determinants influencing procurement and consumption of nutritious diets. This novel empirically grounded conceptual framework can guide future research priorities, analytical approaches, and key intervention points to optimise women’s nutrition.

## 1. Introduction

There is a paucity of evidence to guide transformative change within food systems to support human and planetary health,^1^ especially for the most vulnerable^2,3^ in low-resource contexts.^4–8^ Rapidly changing food environments are associated with malnutrition in all its forms,^9,10^ including persistent micronutrient deficiencies,^11^ and an exponential increase in diet-related non-communicable diseases.^12^ The Lancet’s series on malnutrition, sustainable food systems, social equity, and maternal nutrition and health highlight a long-standing emphasis on biomedical approaches. Limited attention has been given to human rights^13^ and preventative food-based approaches^10^ to address broader eco-social determinants of health,^1,14^ especially for women.^15^ Dual-purpose actions are needed to address determinants of malnutrition in all its forms through cost-effective systems-based solutions^16^ which address unsustainable food system and dietary practices,^1^ persistent gender disparities,^2^ and the intensifying climate-change and malnutrition crisis.^14^

Women of reproductive age (WRA) in low- and middle-income countries (LMICs) are disproportionately affected by malnutrition, compared with men,^14,17^ associated with heightened nutrition needs and gender inequality.^18–21^ Globally, over two-thirds of women lack access to healthy diets, with 1.2 billion experiencing one or more micronutrient deficiencies.^11^ Less than 20% of pregnant women in LMICs meet recommended nutrient intakes.^22^ Despite ambitious global targets, crises such as pandemics, climate extremes, conflict, and economic downturns contribute to growing inequalities, perpetuating and deepening food insecurity and malnutrition of women.^17^ No country is on track to meet the Sustainable Development Goals (SDG) for maternal nutrition.^17^

Food environments - where individuals acquire food from the wider food system - are a key entry point to improve nutrition.^3^ Multiple conceptual frameworks for food environments have been developed^4–6,23–25^ ranging from the globally applicable^5,23^ to specific contexts^6,24,25^ and populations.^4^ However, current frameworks are not gender-sensitive, risking the effectiveness of nutrition initiatives.^16,26–28^ There is growing recognition of the need to consider the interaction of influences across multiple levels and determinants of the external (national, regional, community, institutional) and personal (social, household, individual) food environment dimensions.^4,6,23,25^ There is also a need to incorporate food security elements,^23^ including new pillars of agency and sustainability.^13^ Most empirical research, which applies food environment frameworks, has been on urban markets in high- and middle-income countries.^6–8,14,25^ Little is known about food environments in low-income countries and rural/peri-urban areas where the double burden of malnutrition is intensifying alongside scarce health resources.^9^

This study aimed to develop the first empirically grounded food environment conceptual framework for women in LMICs by employing a systematic scoping review to identify determinants of food acquisition and dietary intakes. By mapping key determinants, this study will guide effective actions to improve women’s nutrition, health and wellbeing through equity and sustainability lenses.^3,13^

## 2. Methods

### 2.1 Study design

The protocol was pre-published.^29^ Briefly, a systematic scoping review was conducted to synthesise evidence from heterogenous disciplines to map the evidence base and define concepts,^30^ according with methodology by PRISMA-ScR^31^ and Joanna Briggs Institute.^32^ The PRISMA-ScR checklist and expanded methods are outlined in the Supplementary Information (SI) (pp.2-6).

### 2.2 Search strategy and study selection

This review considered peer-reviewed quantitative, qualitative, mixed method, or review studies published in English, Spanish, Portuguese, or French between 2010 and 2023 reporting on influences of ≥1 food environment determinants on ≥1 food acquisition practices and/or dietary intakes of WRA (aged 15-49 years) in LMICs, as defined by the World Bank in 2021 (**Table 1**). Databases (*n*=21) across EBSCO, Web of Science Core Collection, and PubMed were searched using a broad criterion based on emerging food environment concepts,^5,23^ and the expanded food security definition.^13^ The final search was conducted on May 26, 2023. Screening was performed in duplicate by researchers fluent in the relevant language, with discrepancies resolved by group discussion. The PRISMA flow chart reports the study selection process (**Figure 1**). From 3,751 retrieved studies, 518 were included. The most common reasons for exclusions were data were not disaggregated by gender, outside the target age, or household/child-level outcomes.

**Table 1:** Inclusion and exclusion criteria according to participants, concept, and context and types of studies.

**Figure 1:**
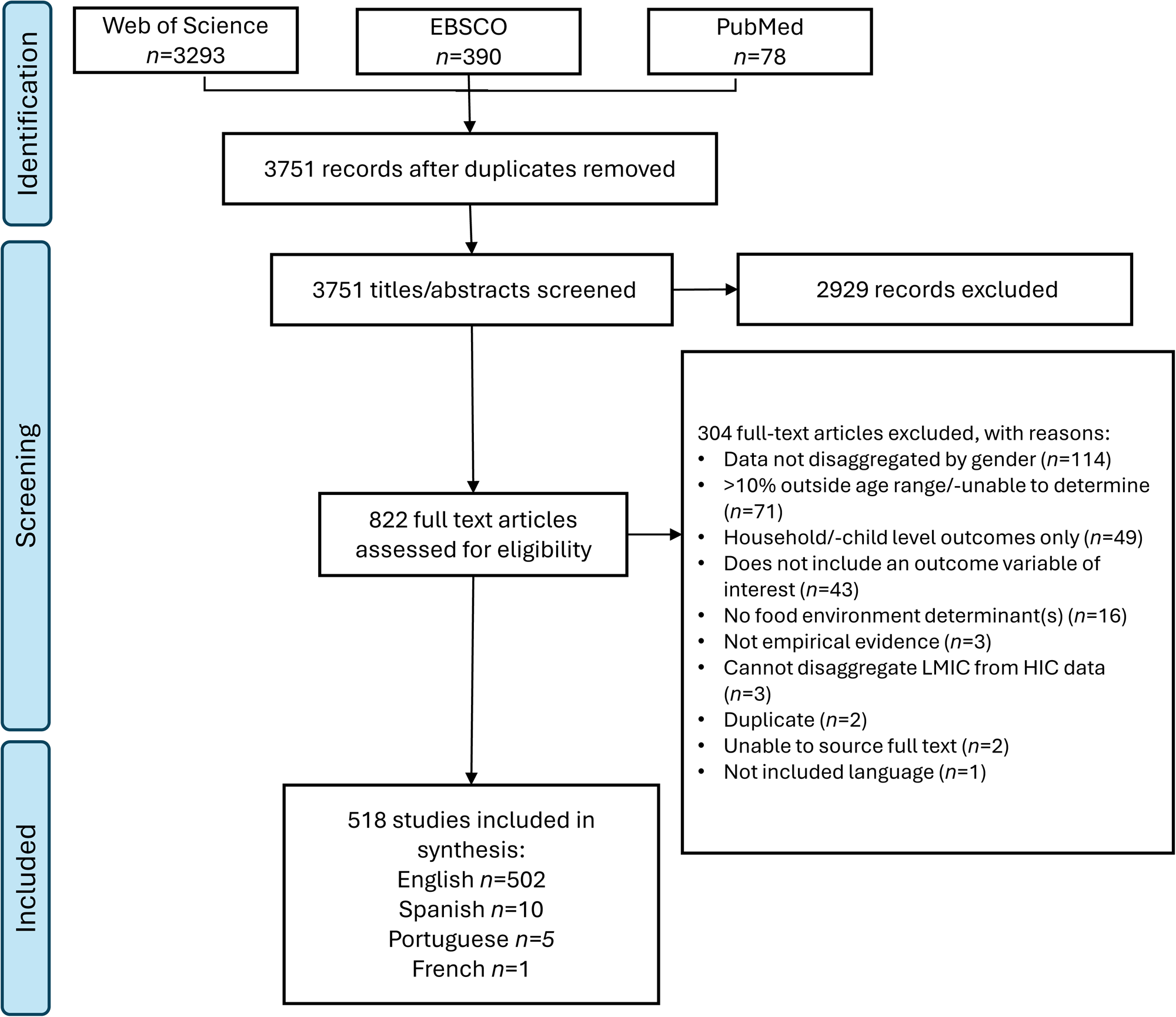
Prisma flow chart of included studies. Articles were published between 1 January 2010 and 30 April 2023.

### 2.3 Data extraction and synthesis, and framework development

Data was extracted on participants, concept, context, study design, methods, and findings relevant to the study objective. Determinants were inductively extracted using a tool iteratively developed by the team. Due to resource constraints, data was extracted singularly. The patterns of outcomes were mapped against identified determinants and existing frameworks.^4–6,23–25^ A preliminary framework was presented at two international academic conferences in 2023. Iterative discussion amongst authors led to the final selection of key determinants in our framework. Given the large volume of material, only the most relevant publications are referenced throughout the results; all included studies are in the appendix (**SI Table 4**).

## 3. Results

### 3.1 Descriptive statistics

Our review included 518 studies, representing 125 countries (**Figure 2; SI Table 2**). Most reported determinants of food acquisition and dietary intakes of non-pregnant, non-lactating (NPNL) women (or unspecified physiological status) (73.6%), with fewer on pregnant (30.5%) and lactating women (16.0%) (**Table 2**). Most were in sub-Saharan Africa (SSA) (41.1%) and South Asia (SA) (20.7%), spanning lower-middle (44.6%), upper-middle (24.9%), and low-income (22.2%) countries. Rural (70.3%) and urban/-peri-urban (55.4%) contexts were represented. Most employed only quantitative methods (62.4%). Dietary intake as an outcome variable was more often considered (94.2%) than food acquisition (47.9%). Annual publication numbers increased from 2010, with exponential growth from 2016 onwards (**SI Figure 1**).

**Figure 2:**
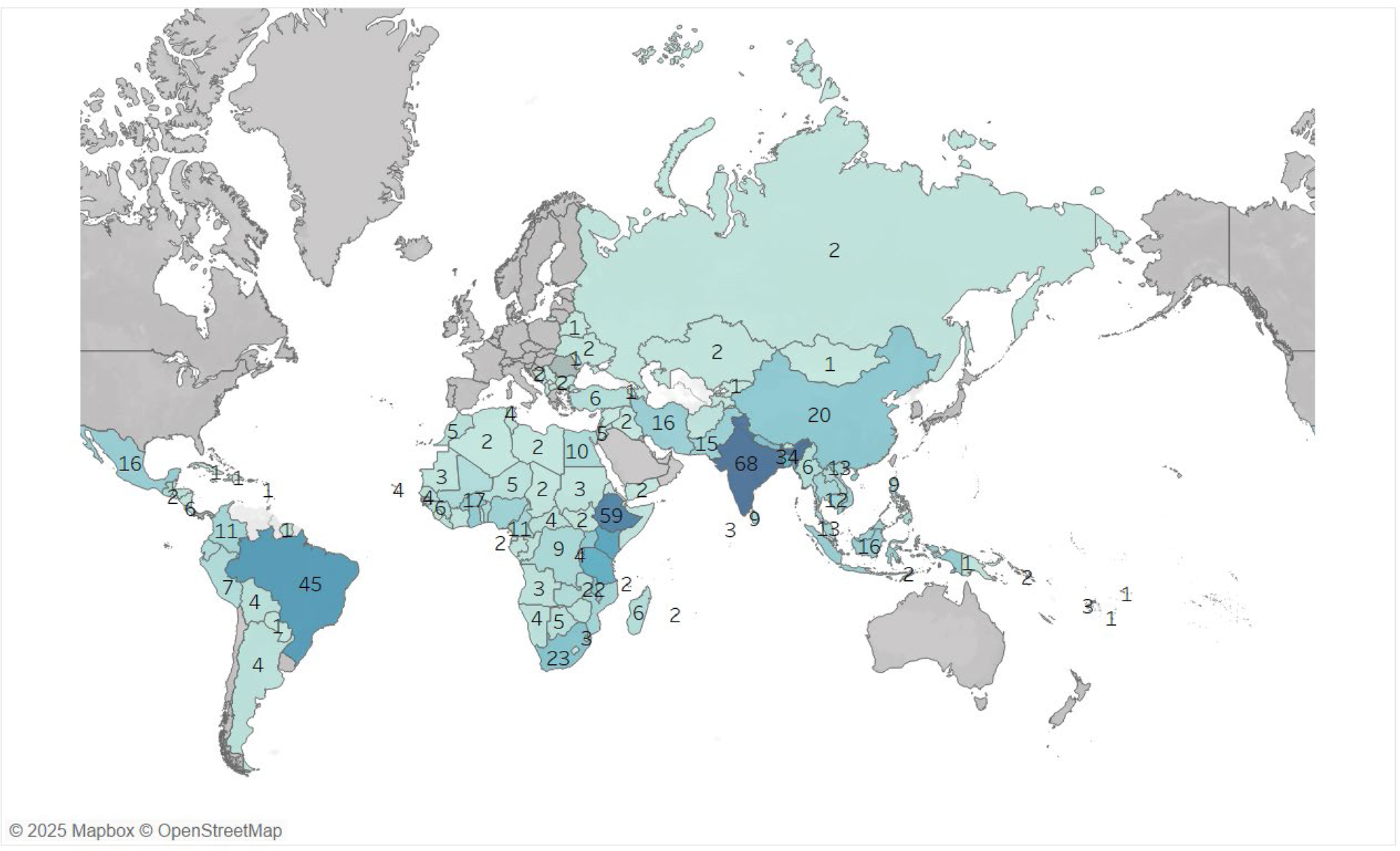
Map demonstrating the distribution of included studies (n=518) by country (n=125). See SI Table 1 for full details. Some of the multi-country studies were not included in the above visualisation because the countries could not be identified from the text.^33–38^

**Table 2:**
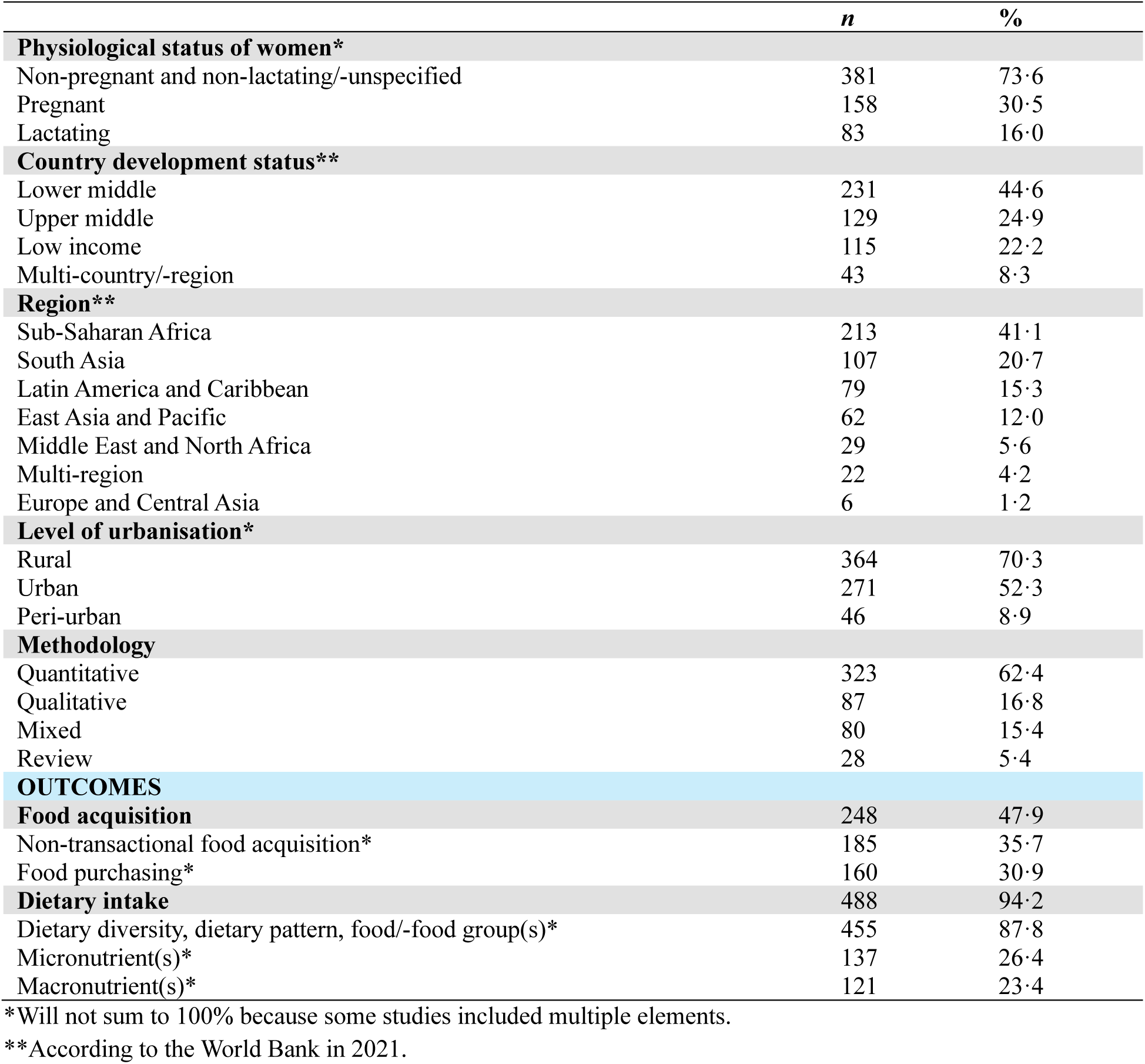
Descriptive statistics of included studies (n=518).

### 3.2 Conceptual framework

The conceptual framework (**Figure 3**) distinguishes four socioecological layers of influence on women’s food acquisition and dietary intakes, building on prior frameworks.^4–6,23,25^ External determinants are reflected at national, regional, community, and institutional levels, while personal determinants exist at household and individual levels, reflecting wider social contexts. We include a novel layer: women’s agency, characterised by an individual’s ability to exert control over resources, as a key mediator of women’s food acquisition and dietary intakes. Sustainability and stability are represented in the outer ring as critical elements across all levels and determinants. Fifteen key food environment determinants were identified across regions (**Figure 4**), informed by 143 sub-themes (**Table 3**; **SI Table 4**). A third of studies reported on women’s agency (35.9%), and stability and sustainability (32.6%), with a quarter on food security (22.8%). Food security is represented in the framework by its ‘pillars’: availability, accessibility (physical, financial, and social), stability, agency, and sustainability. The most reported determinants were food-availability (49.8%), literacy (57.1%) and affordability (52.9%) (**Figure 4; SI Table 2 & 4**).

**Figure 3:**
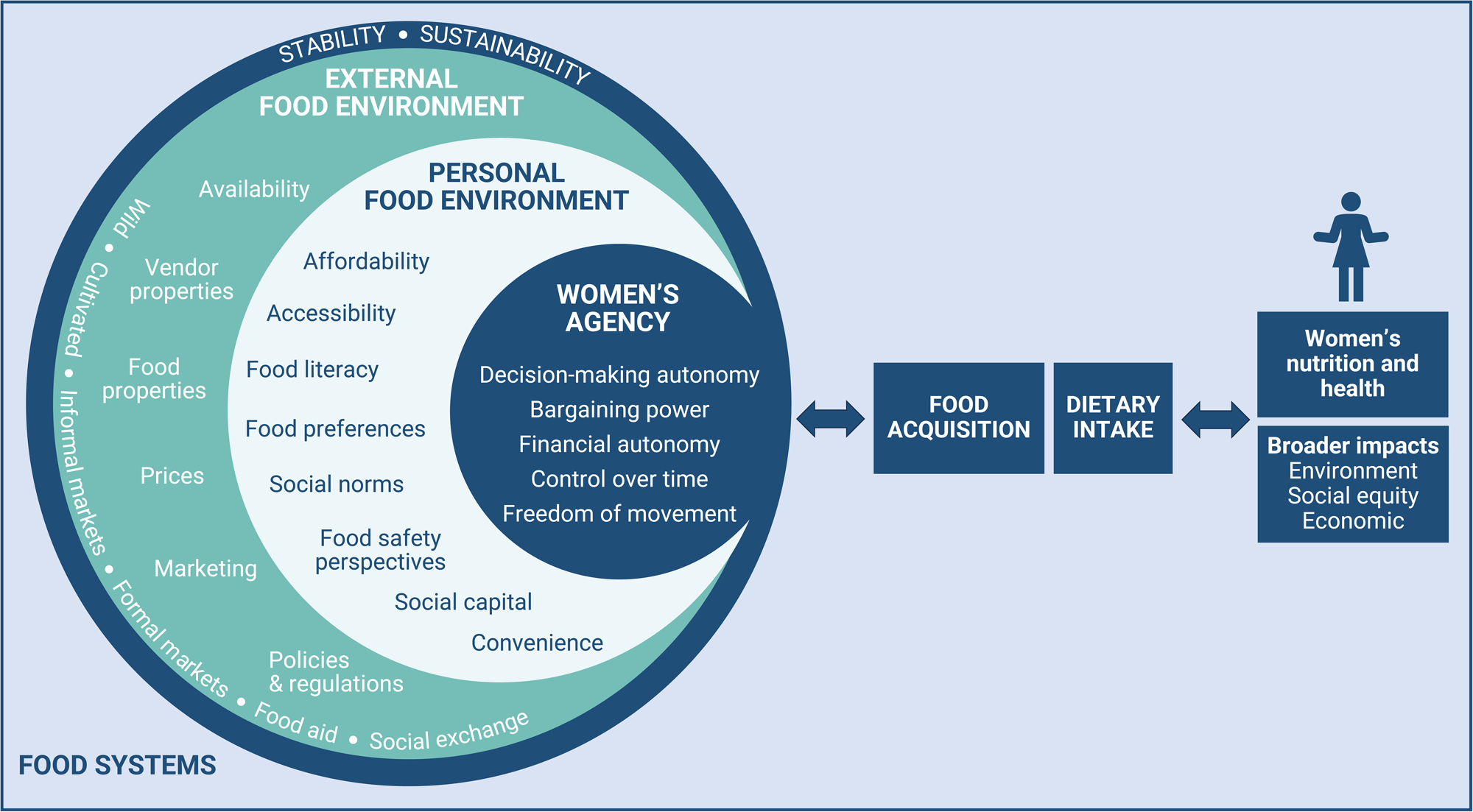
Conceptual framework of women’s food environments, presenting key determinants of food acquisition practices and women’s dietary intakes in LMICs derived from 143 themes identified from the literature. The external food environment presents determinants at the national, regional, community, and institutional levels, whilst the personal environment highlights social, household and individual determinants. A novel layer is presented: women’s agency as a key mediator for the ability of an individual to exert control over resources contributing to one’s own dietary outcomes. Stability and sustainability are key considerations across all levels and determinants. Source: Authors’ own. LMICs = low- and middle-income countries.

**Figure 4:**
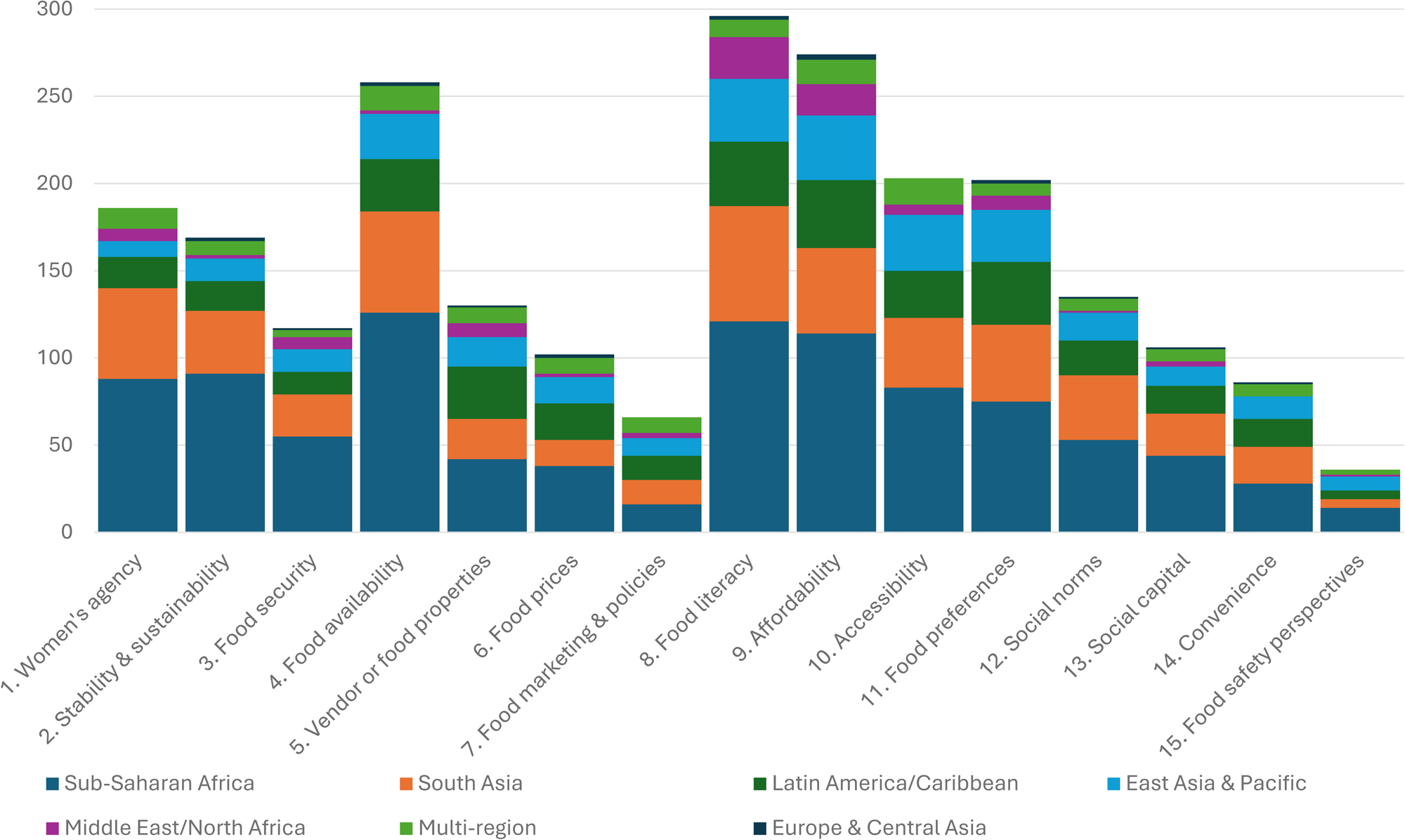
Number of included papers (n=518) reporting on each key determinant (n=15) of food acquisition practices and dietary intakes for women in LMICs, overall and by geographic region. See SI Table 2 for full details.

**Table 3:**
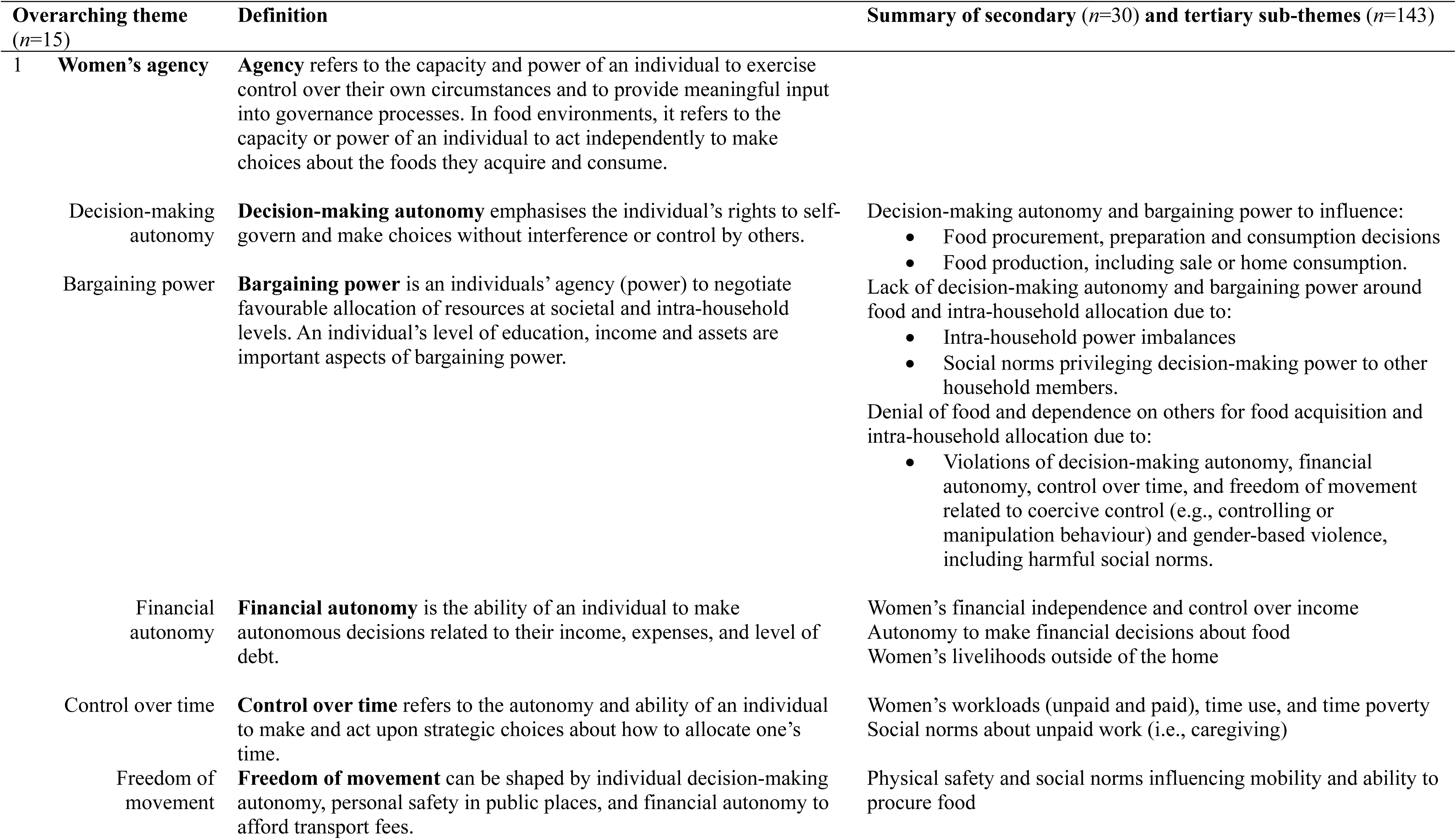
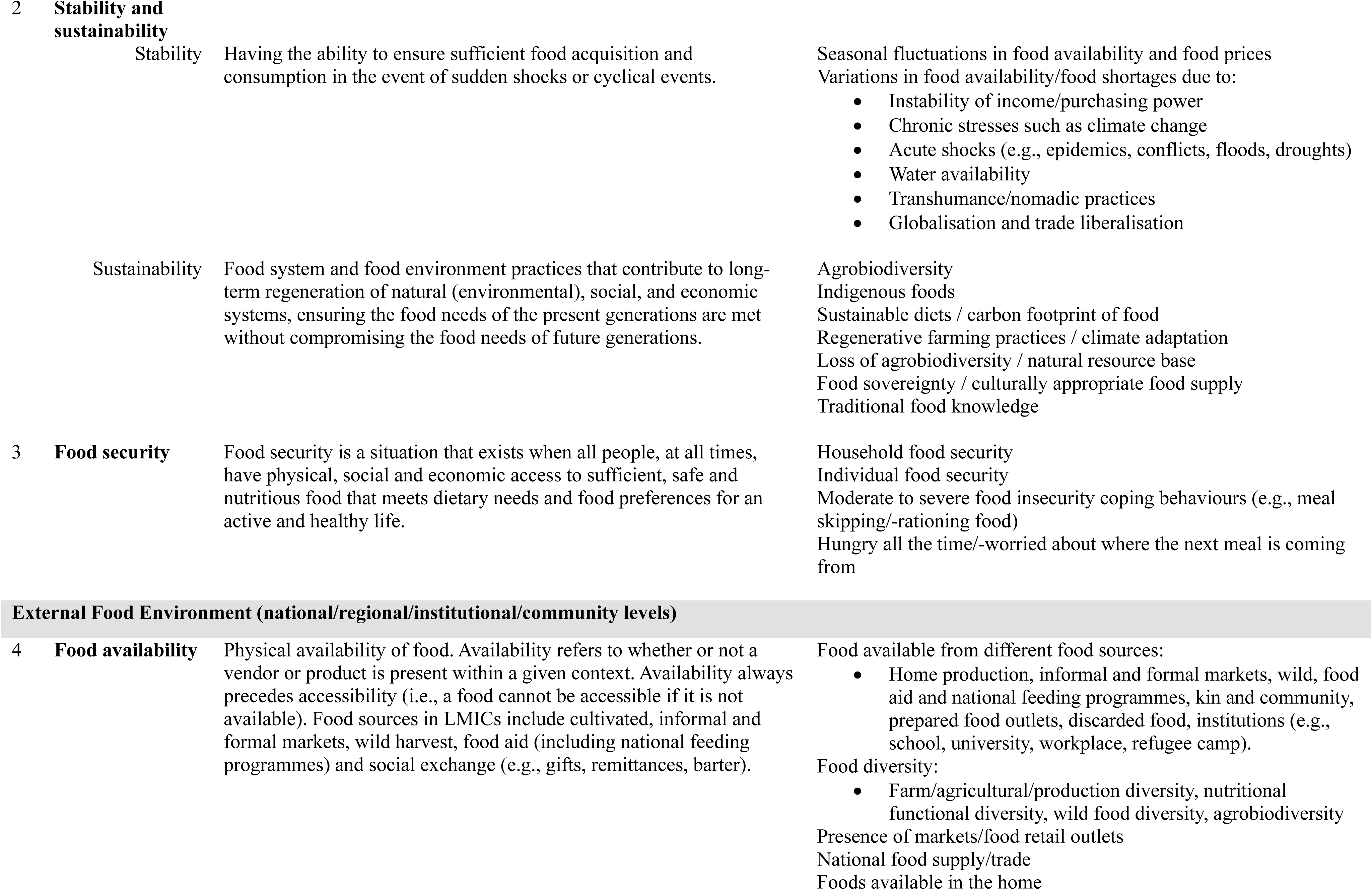
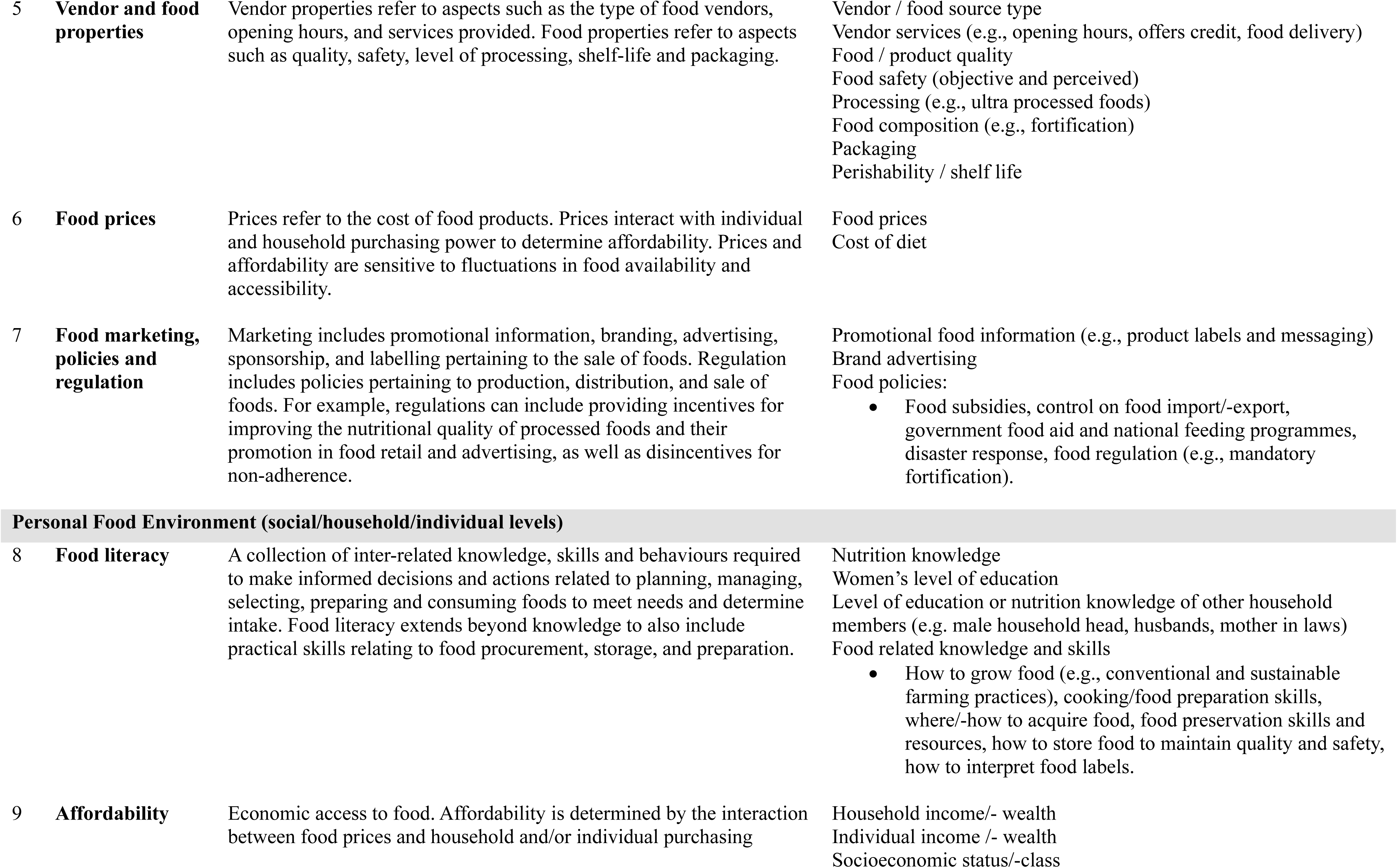
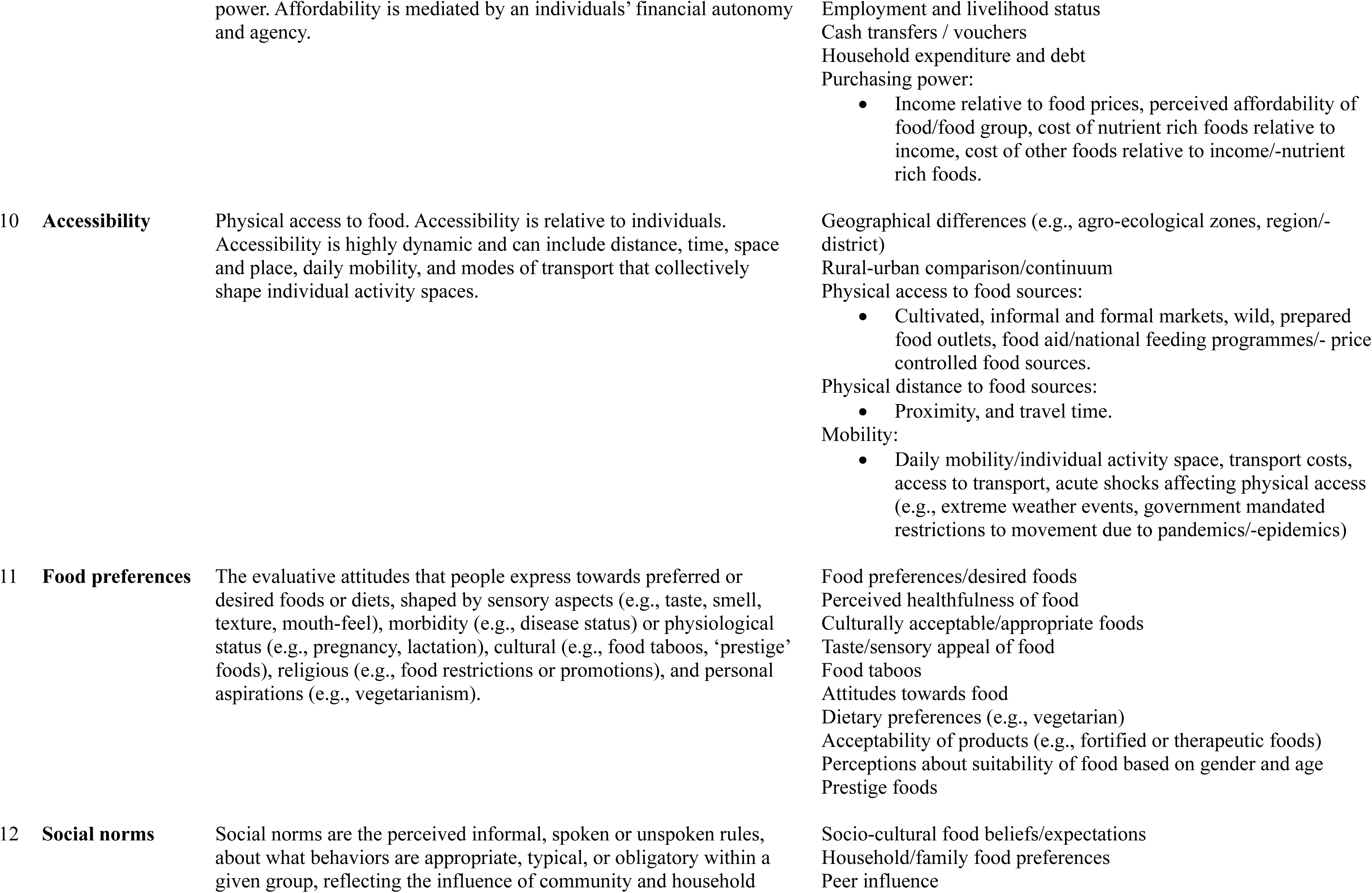
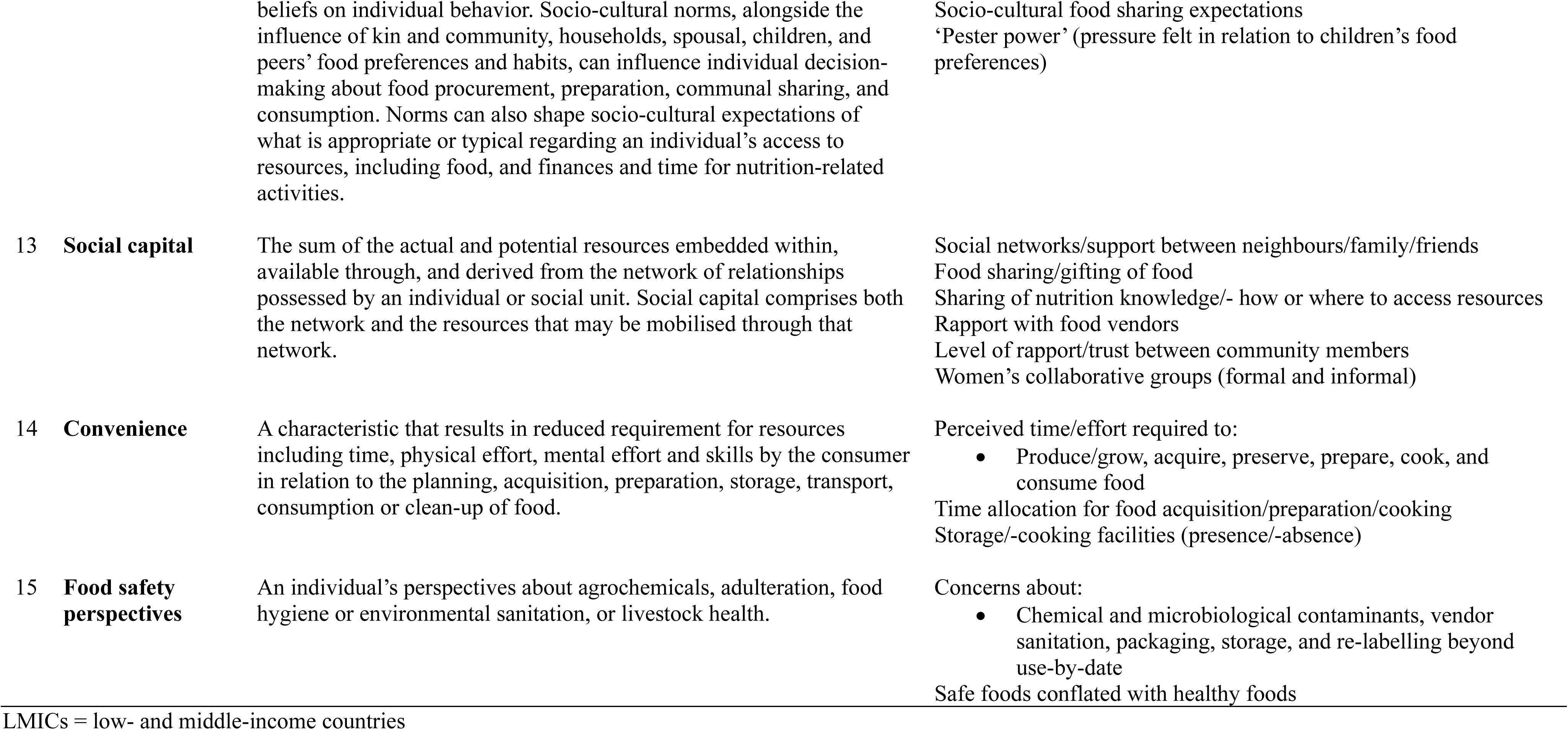
Definitions of the overarching themes and sub-themes, for key determinants of food acquisition practices and dietary intakes of women in LMICs, presented in our conceptual framework. These themes and sub-themes were identified through the literature search (n=518 papers reviewed). See SI Table 3 for full glossary of terms and SI Table 4 for full list of sub-themes and corresponding references.

### 3.3 Women’s agency

#### 3.3.1 Decision-making autonomy, bargaining power, and financial autonomy

Social norms shaped women’s agency in making dietary decisions. Lack of autonomy over food-related decisions, including finances, mediated women’s dietary quality in the Asia-Pacific,^35,39–48^ and SSA.^49–55^ Men often controlled animal-source food (ASF) purchases, especially meat,^40,44,50,51,54–56^ leading to women consuming less iron-rich foods.^46,50,54–56^ Women often bought food, based on husband and children’s preferences^57–60^ or as decided by male household heads.^47,50,51^ Men also dominated decisions around food production, and sales.^40,49,53,61–63^ Social norms prioritising food allocation to other household members affected women’s dietary quality in SA,^43^ and SSA.^52^ Within households, women often ate last.^35,40,43,45,50,51,53,55,64–66^ It was socially expected that women prioritise the food needs of other household members, a norm sometimes enforced by gender-based violence,^39,45^ with subsequent negative effects on women’s dietary quality compounded by food insecurity.^40,43,45,50,52,55,57,64,66–71^ Preferred meat cuts and larger quantities were often privileged to men; while there were instances of milk and eggs given preferentially to children,^40,51^ with inequitable intra-household allocation of ASF linked with women’s lower dietary quality.^40,43,45,50,53,64^

Urbanisation tempering gendered food provisioning in SA,^43,46^ and SSA,^59,72^ with examples of men and other household members sharing unpaid workloads and purchasing additional foods for pregnant women.^59,63,67,73^ Examples of more equitable food allocation were found in some contexts.^40,74,75^ However, in SA, the dietary diversity score (DDS) gap between genders was high,^76^ often regardless of household socioeconomic status or caste.^43,46^ Coercive control, manipulative behaviours, economic abuse, and physical and sexual violence constrained women’s control over food.^39,40,45,50,77^ Women were sometimes recipients of violence if actions around food were perceived to question existing power dynamics, such as allocating equal portion sizes;^45^ contributing to decision-making and financial processes on food,^45,61,77^ including purchasing meat themselves^40,50^ or changing foods cooked.^39^ Withholding food was an extension of violence by perpetrators,^45^ with some women consuming less or lower quality foods in abusive contexts.^40,45^

#### 3.3.2 Control over time and freedom of movement

Women carried disproportionately higher work burdens due to unpaid caregiving work.^43,55,56,64,78^ Where women had limited control over time, paid employment^43,58,79–83^ and agricultural work ‘doubled’ work burdens.^62,63,77,78,84^ Time poverty negatively affected women’s dietary diversity scores (WDDS) in SSA and Asia-Pacific,^78^ driving convenient food consumption (section 3.6.3). During planting and harvesting periods, pregnant women sometimes ate less due to lack of time and exhaustion.^43,67,70^

Social norms and physical safety concerns restricted women’s mobility, limiting food access globally,^36,85^ including SSA,^72,73,85–88^ Asia-Pacific,^85,89^ and Latin America/Caribbean (LAC).^85^ Social norms restricting mobility were strong in SA,^36,46,47,85^ where food markets were considered ‘male’ spaces.^46,47,56,90^ There were incidences of more freedom of movement in SSA,^91^ and SE Asia.^42^

### 3.4 Stability, and sustainability

#### 3.4.1 Stability

Instability in physical and economic access to food negatively affected women’s dietary intake,^92,93^ especially in SSA,^86,92–99^ Asia-Pacific,^42,89,92,93,100–108^ LAC,^92,93,109–112^ and Middle East/North Africa (MENA).^92,113^ Longitudinal studies demonstrated dietary quality changes across seasons,^42,64,94,96–99,101,102,114^ affecting WDDS,^64,97,98,101,102,114^ and women’s nutrient intakes.^94,96,99^ Protracted conflict- and climate-related crises,^99,115–119^ acute shocks,^86,89,92,93,104–113^ and severe weather events^95^ impacted women’s dietary quality. The socio-ecological shocks of the Covid-19 pandemic impaired affordability and accessibility of food globally,^92^ deteriorating women’s dietary quality in Asia,^105–107^ Africa,^86,113^ and Small Island Developing States (SIDs),^93^ especially for adolescent girls, and pregnant and lactating women in the most resource-constrained contexts with severe pandemic-related restrictions.^86,89,104^ During food security crises, less negative dietary effects were noted for women receiving food aid^95,107,109^ or who had more access to cultivated or wild foods.^106,110^ During the pandemic, there were examples of women’s dietary quality deteriorating more than men,^113^ and national feeding centre closures were correlated with WDDS declines.^105^ In times of severe food insecurity, some women ate unsafe or low-quality foods.^68,86,120^ Women’s dietary quality also suffered due to maternal buffering, where women protected food for children during food shortages in SSA,^51,55,64,67,70,120^ Asia-Pacific,^40,42,45,90,107,121,122^ and LAC.^68,71,110,123^ During food insecurity periods, some food-sourcing strategies included survival sex,^86,120^ and scavenging discarded food.^68,120^

#### 3.4.2 Sustainability

Food biodiversity correlated with improved dietary quality across seasons and the rural-urban continuum in SSA,^124–129^ LAC,^130–132^ Asia-Pacific,^76,103,133^ and MENA.^134^ Agrobiodiversity of cultivated and wild food species correlated positively with diversity of WDDS/MDD-W,^125–127,129,131^ and micronutrient intakes/adequacy,^76,129,131,132^ including for diversity of indigenous foods consumed.^132,135–139^ A few studies investigated indigenous food systems and transformative farming practices in SSA,^73,129,140,141^ LAC,^61,62,130^ and Asia-Pacific.^75,103^ Degradation of natural resources limited wild and cultivated food supplies for marginalised groups.^73,75,100,103,122,140,142^ Whereas agroecological practices were associated with improved food sovereignty,^61^ production diversity, WDDS, and micronutrient intakes.^129,130^ Agrobiodiversity, especially of indigenous foods, was valued by women’s ethno-nutrition knowledge as climate-adaptive strategies for overcoming seasonal variations in food availability.^73,141^ Some women faced gendered barriers to adopting transformative farming practices,^61,62,73,140^ including violent community ‘backlash’,^61^ double work burdens, lack of land or agricultural asset ownership, and constrained decision-making autonomy.^62,140^ Limited studies analysed sustainability of current diets.^143–146^ Food costs were a barrier to adherence to sustainable diets for lactating women from food-insecure, rural contexts in SSA,^146^ whereas socio-cultural traditions and convenience were barriers in urban LAC.^144^

### 3.5 External food environment

#### 3.5.1 Availability

Women acquired food from a diversity of sources. Across the rural-urban continuum, fresh produce markets (predominately informal) were a dominant source of unprocessed, nutrient-rich foods in SSA,^80,87,92,125^ Asia-Pacific,^85,92,133,147^ LAC,^92,130,148^ and SIDs.^93^ In rural and peri-urban areas, home production and wild environments were important healthy food sources^54,76,84,85,92,93,100,106,121,127,128,130,135,139,149–151^ as were informal vendors and fresh produce markets in urban areas,^87,92,133,147,148^ and non-agricultural rural contexts.^58,80^ Food aid and national feeding programmes were important food sources globally,^37^ especially in SA,^105,107,136,152^ SSA,^67,118^ and LAC.^109,123^ Social exchange of food was common (section 3.6.4).^40,42,45,46,51,53,59,60,67,68,120–122,130,153,154^

#### 3.5.2 Vendor and food properties

In SSA, microbial and aflatoxin spoilage were key food safety risks,^35,51,72,85,87,155,156^ compared with chemical contamination in Asia-Pacific.^57,60,147,155,157–162^ Ultra-processed foods were a common concern in LAC,^22,85,109,111,112,130,163–165^ and SIDs,^100,103^ associated with inadequate micronutrient intake among pregnant women in LAC.^163–165^ Concerns about vendor cleanliness, adulteration, and mislabelling also drove vendor and food choices.^51,57–60,80,85,87,147,160^ Women valued vendors that provided credit and sold fresh, high quality, affordable foods.^51,58,60,80,87,147,162,166^ Some purchased direct from farmers for reduced costs and higher quality.^51,60,87,157,162^

#### 3.5.3 Food prices

Cost of a nutritious diet was a challenge globally,^167^ especially in SSA,^146,167,168^ Asia-Pacific,^167,169^ and MENA.^167,170^ High prices for ASFs and vegetables drove costs.^146,167–169^ Although realistic diets modelled on low cost, locally available and culturally appropriate foods could provide most recommended nutrient intakes; it was not always possible to achieve adequacies for all micronutrients due to the cost of meat.^35,169,170^ Iron was the most expensive micronutrient, especially to meet needs during menstruation and pregnancy.^35,169–171^ Although women aspired to purchase natural or unadulterated foods, price was a barrier.^57,60,87,147,157,162^

#### 3.5.4 Marketing, policies and regulation

Marketing, alongside attractive packaging and children’s ‘pester power’ (section 3.6.2), were linked with processed food purchases.^33,57,60,72,75,82,85,87,88,147,155,160,162,172,173^ Food marketing was more common in urban areas.^82,87,147,155,174^ Nutrition labels had potential to influence food purchases;^175–179^ however, technicality and cultural incompatibility of information, and lack of trust in manufacturers limited effectiveness.^175–177^

Most regulations involved fortification of staple foods with micronutrients;^22,55,123,154,180–189^ however, there was insufficient evidence to determine effectiveness for women.^38,154,180,184,188,190–192^ Some evidence of urban policies privileging supermarkets was correlated with higher produce costs in SE Asia.^60,133^ Few studies existed on the effects of agricultural policies, biofortification, globalisation, international trade, and food subsidies on women’s dietary intakes.^38,193,194^

### 3.6 Personal food environment

#### 3.6.1 Affordability

Low purchasing power constrained nutrient-rich food purchases globally,^35,36,85,92,93,155^ including in SSA,^33,50,51,67,70,72,86–88,99,128,195^ Asia-Pacific,^42,43,76,89,100,105,106,121,122,147,162,196–199^ LAC,^68,82,83,109,110,144,200^ and MENA,^116,201,202^ especially during shocks.^86,89,92,93,95,99,105,106,109,110,115–117,119,121^ Economic constraints were key barriers to dietary adequacy during pregnancy and lactation, especially for ASF.^52,85^

#### 3.6.2 Food literacy, preferences, perceptions, and social norms

Women’s food literacy, food preferences, and social norms mediated pathways between financial and physical access with dietary quality in SSA,^33,49,52,67,72,85,87,88,195,203^ Asia-Pacific,^42,85,100,147,204,205^ and LAC.^82,83,85,200,206^ The food literacy and preferences of household decision-makers also mediated pathways between women’s nutrition knowledge and dietary quality when decision-making autonomy was low in SA,^43,44,63,207–210^ SSA,^64,70,73,211,212^ SE Asia,^40,213^ LAC,^61,62^ and MENA (section 3.3).^214^ Religious fasting and food taboos often limited women’s consumption of nutrient-rich foods,^35,42,43,52,53,55,64,65,67,70,155,212,215,216^ correlating with lower dietary quality.^43,154,198^ Family members were key influencers of dietary intake during pregnancy and lactation,^43,52,53,67,70,88,212^ while peer pressure influenced adolescent girls’ food choices.^59,85,160,173^ In LAC, children’s ‘pester power’ drove ultra-processed food purchases and consumption by mothers.^81,83,217^ Food safety was sometimes conflated with healthy food.^57,80,85^

#### 3.6.3 Accessibility and convenience

Accessibility mediated food acquisition globally,^85,92,93^ especially in SSA,^67,72,87,88^ Asia-Pacific,^42,60,147,159^ and LAC.^148,200^ Purchasing food in bulk at large produce markets or supermarkets was perceived as cheaper but not always possible due to time, financial, and cold storage constraints,^58,60,72,80,85,87,88,148^ correlated with reduced odds of nutrient-rich food consumption.^159^ Time constraints were linked with increased demand for convenience foods in LAC,^79,81–83,144,217,218^ SSA,^58,72,80,87,155^ and Asia-Pacific.^57,60,155,174,177,219^ Easier access to unhealthy foods was linked with higher presence of food vendors,^33,79,82,83,85,88,100,145,147,148,174,177,202,217,220,221^ especially near schools and workplaces.^79,85,88,144^ Access to healthier foods was higher in places dominated by informal vendors close to homes.^60,147,148,151,159^ Poor infrastructure for walking and personal safety concerns limited access to urban food markets;^85,87,88^ whereas distance and social norms were stronger constraints rurally (section 3.3).^67,76,77,222^. Pandemic-related mobility restrictions,^86,89,93,105,106^ and extreme weather^95^ also limited access.

#### 3.6.4 Social capital

Social networks were important sources of food. In times of financial hardship, women often bought food on credit from known vendors^42,50,51,58,68,80,87^ or sourced food or money from kin and community.^42,45,46,51,59,60,68,120–122,130,153^ In low-resource settings, cultural ceremonies were an opportunity for ASF consumption.^40,53,67,120,154^ Women leveraged social networks to divide unpaid labour demands,^58,60,81,153,223,224^ and collaborative buying power for bulk food purchases.^60,166^ Women’s groups provided access to food- and agriculture-related knowledge and resources, and food exchange.^36,44,45,63,118,130,152,203,225^

## 4. Discussion

This paper presents the first gender-sensitive conceptual food environment framework for women in LMICs, empirically grounded in 143 inductively identified determinants of women’s food acquisition practices and dietary intakes from 518 studies across 125 countries. The inclusion of novel determinants, including integration of gender disaggregated evidence on agency, sustainability, and food security, expands existing food environment frameworks, and highlights key considerations for women in LMICs. The framework is representative across multiple contexts, including historically under-represented rural and peri-urban settings in low-income countries, contrasting with previous frameworks and reviews which employed expert consultative^5,23^ or deductive approaches,^7,8^ predominately deriving evidence from higher-income and urban contexts.^6–8,25,33,85,155^

Consideration of women’s agency is critical if policies and programmes aim to improve equality in dietary quality.^14,20,21^ Social norms mediated women’s agency (ability to exert control) over food-related decisions, finances, workloads, and mobility.^26^ This influence, affecting their ability to translate nutrition knowledge into action, aligns with correlations between women’s nutrition empowerment with women’s dietary outcomes and nutrition in LMICs.^19–21^ Initiatives often target women as agents for sustainable food production, food security, and household dietary changes. If such efforts are designed without identifying the relative importance of various determinants – and addressing women’s agency – their effectiveness may be limited,^20,21^ risking inefficient use of resources and unintended gender consequences.^27,226^

Women’s nutrition interventions and research will require a holistic systems approach considering interlinkages between framework levels and distinguishing enabling resources such as nutrition knowledge from agency.^20,21^ A woman may have economic and physical access to nutritious food, but limited agency may constrain procurement and consumption. It is important to differentiate nutritional empowerment from other empowerment forms, such as educational and economic, which may not automatically improve nutrition due to mediators such as lack of decision-making autonomy. Context-specific approaches are essential due to varying facilitators and barriers across geographic, governance, socioeconomic, and cultural determinants.^26^

The mediating effect of agency on uptake of climate-adaptive practices also needs to be considered. The negative results of disempowerment may occur in a feedback loop,^36^ compounding the effects of climate change, unsustainable use of natural resources, food insecurity, conflict and malnutrition.^227,228^

Changes in governance, laws, policies, and infrastructure are necessary to support women’s agency and develop healthier, more resilient enabling food environments.^26,229^ Broader policies around land tenure, agriculture, trade, infrastructure, and social protection influence availability and accessibility of foods at multiple levels, underscoring needs for multi-sectoral approaches.^28^ The pervasive influence of commercial determinants of consumption requires sustained focus on developing effective strategies for targeting external food environment determinants, including healthier food costs and regulation of ultra-processed food availability and marketing.^14^ While biofortification of crops shows potential for addressing women’s micronutrient deficiencies, studies need to assess intermediate indicators that may mediate uptake among women.^38^

Different food sources and biodiversity protected women’s dietary quality, particularly during shocks when women needed to diversify food sources to address food insecurity.^24,87,92,147,230^ Given the importance of biodiversity to resilient ecosystems, more research is needed to understand the contribution of different food sources to sustainable and nutritious diets for women.^1,14^

We found that women’s dietary outcomes were particularly vulnerable to shocks and scarcity, indicating that the historical reliance on household-level indicators is inadequate for assessing women’s diets. Women’s dietary intake could serve as a sentinel indicator for monitoring food security and dietary changes in response to crises and food environment transformations, especially within gender imbalanced contexts. The routine collection of women’s individual level indicators is warranted, including MDD-W in Demographic and Health Surveys, and SDG monitoring.^17^

The literature review found a striking lack of focus on sustainability and external food environment determinants, limiting our understanding of climate change and macro-level policy and programme effects. There was scant evidence on food acquisition practices and quantified micronutrient intakes, limiting our ability to identify leverage points for addressing micronutrient deficiencies in women.^17^ Although there was a noticeable shift towards systems approaches after publication of leading frameworks on food environments,^5,23^ there is still a lack of holistic, multi-disciplinary studies considering feedback loops and trade-offs. The bulk of evidence was small-scale, quantitative, cross-sectional studies, with a lack of standardised methods for assessing determinants across long causal pathways, limiting comparability across studies. The smaller number of qualitative studies contributed insights into women’s lived experience, although there was a reliance on perceived rather than objective measures of some determinants such as food safety. There was a noticeable lack of focus on adolescent girls and lactating women, and women in climate- and conflict-affected regions.

Exclusion of grey literature is a limitation of this review; however, inclusion of articles published in four languages with only a small number identified in Spanish, French, and Portuguese indicates high coverage. Although search terms used may have inadvertently excluded relevant studies, broad search criteria enabled synthesis of heterogenous evidence across multiple disciplines and geographies. Some overlap of concepts was inevitable when using a socio-ecological framework, yet it has advantages for identifying policy entry points.

## 4. Conclusion

We present the first conceptual framework of women’s food environments in LMICs, empirically grounded in a review of 518 studies from 125 countries. Sustainable approaches to improving women’s nutrition requires policies and programmes address underlying legislative, structural and socio-cultural determinants mediating women’s agency, alongside other determinants influencing nutritious diets. By addressing key determinants through coordinated and targeted actions, policymakers in LMICs may improve the effectiveness of nutrition interventions, particularly where changing environmental, political, and economic contexts threaten to perpetuate nutrition inequalities. Women’s dietary intake is often differentially affected compared to other household members, warranting individual-level monitoring. While the interrelated effects of determinants are complex, the lack of progress on women’s nutrition underscores the need for multi-sectoral actions to improve women’s agency and food environments. By systematically mapping critical determinants influencing women’s food acquisition and dietary intakes, we identified novel food environment determinants, resulting in a robust framework applicable across LMICs and the rural-urban continuum. This framework can guide future research priorities, analytical approaches, and key intervention points to optimise women’s nutrition.

## Data Availability

A searchable database of the reviewed studies is available at:
O'Meara, Lydia (2025), 'Systematic scoping review: Determinants of women's food acquisition practices and dietary intakes in LMICs', Mendeley Data, V1, doi:10.17632/x4rcx7bmdx.1

https://data.mendeley.com/datasets/x4rcx7bmdx/1

## Data sharing

A searchable database of the reviewed studies is available at: O’Meara, Lydia (2025), “Systematic scoping review: Determinants of women’s food acquisition practices and dietary intakes in LMICs”, Mendeley Data, V1, doi:10.17632/x4rcx7bmdx.1

## Author contributions

Conceptualisation (LO, JdB, KW, EF, PDS); Data curation (LO, TH, RH, MFP, PDS); Formal analysis (LO, JdB, TH, PDS, MFP, RH, CT, KW, EF); Funding acquisition (LO, JdB, KW); Investigation (LO); Methodology (LO, JdB, PDS, KW, EF, CT, TH, MS); Project administration (LO); Resources (LO, MS); Software (LO); Supervision (LO, JdB, PDS, KW, EF); Visualisation (LO, JdB); Writing - original draft (LO); and Writing - review & editing (LO, JdB, PDS, EF, MFP, CT, TH, RH, KW).

## Acknowledgements

The authors would like to thank colleagues who commented on early drafts. They would also like to thank three third year nutrition and public health students that assisted with early title and abstract screening.

## Ethical approval

This study was based on publicly available literature. It did not include any animal or human participants. Ethical approval and consent to participate was not applicable.

