## Supplementary Information for "Conceptual framework of women’s food environments and determinants of food acquisition and dietary intakes in low- and middle-income countries: a systematic scoping review"

O'Meara et al.

1.PRISMA-ScR checklist

**Preferred Reporting Items for Systematic reviews and Meta-Analyses extension for Scoping Reviews (PRISMA-ScR) Checklist**

| SECTION | ITEM | PRISMA-ScR CHECKLIST ITEM | REPORTED ON PAGE # |
|---|---|---|---|
| **TITLE** | | | |
| Title | 1 | Identify the report as a scoping review. | 1 |
| **ABSTRACT** | | | |
| Structured summary | 2 | Provide a structured summary that includes (as applicable): background, objectives, eligibility criteria, sources of evidence, charting methods, results, and conclusions that relate to the review questions and objectives. | 2 |
| **INTRODUCTION** | | | |
| Rationale | 3 | Describe the rationale for the review in the context of what is already known. Explain why the review questions/objectives lend themselves to a scoping review approach. | 4 |
| Objectives | 4 | Provide an explicit statement of the questions and objectives being addressed with reference to their key elements (e.g., population or participants, concepts, and context) or other relevant key elements used to conceptualize the review questions and/or objectives. | 4 |
| **METHODS** | | | |
| Protocol and registration | 5 | Indicate whether a review protocol exists; state if and where it can be accessed (e.g., a Web address); and if available, provide registration information, including the registration number. | Published protocol: DOI: 10.11124/JBIES-22-00299 |
| Eligibility criteria | 6 | Specify characteristics of the sources of evidence used as eligibility criteria (e.g., years considered, language, and publication status), and provide a rationale. | 5<br>SI pp. 4-6 |
| Information sources* | 7 | Describe all information sources in the search (e.g., databases with dates of coverage and contact with authors to identify additional sources), as well as the date the most recent search was executed. | 5<br>SI pp. 4-6 |
| Search | 8 | Present the full electronic search strategy for at least 1 database, including any limits used, such that it could be repeated. | Published protocol: DOI: 10.11124/JBIES-22-00299 |
| Selection of sources of evidence† | 9 | State the process for selecting sources of evidence (i.e., screening and eligibility) included in the scoping review. | 5<br>SI pp. 4-6 |
| Data charting process‡ | 10 | Describe the methods of charting data from the included sources of evidence (e.g., calibrated forms or forms that have been tested by the team before their use, and whether data charting was done independently or in duplicate) and any processes for obtaining and confirming data from investigators. | 5<br>SI pp. 4-6 |
| Data items | 11 | List and define all variables for which data were sought and any assumptions and simplifications made. | 5<br>SI pp. 4-6 |
| Critical appraisal of individual sources of evidence§ | 12 | If done, provide a rationale for conducting a critical appraisal of included sources of evidence; describe the methods used and how this information was used in any data synthesis (if appropriate). | NA |

| SECTION | ITEM | PRISMA-ScR CHECKLIST ITEM | REPORTED ON PAGE # |
|---|---|---|---|
| Synthesis of results | 13 | Describe the methods of handling and summarizing the data that were charted. | 5<br>SI pp. 4-6 |
| **RESULTS** | | | |
| Selection of sources of evidence | 14 | Give numbers of sources of evidence screened, assessed for eligibility, and included in the review, with reasons for exclusions at each stage, ideally using a flow diagram. | 5<br>SI pp. 4-6 |
| Characteristics of sources of evidence | 15 | For each source of evidence, present characteristics for which data were charted and provide the citations. | 5-10,<br>SI pp.6-23 |
| Critical appraisal within sources of evidence | 16 | If done, present data on critical appraisal of included sources of evidence (see item 12). | NA |
| Results of individual sources of evidence | 17 | For each included source of evidence, present the relevant data that were charted that relate to the review questions and objectives. | 5-10,<br>SI pp.6-23 |
| Synthesis of results | 18 | Summarize and/or present the charting results as they relate to the review questions and objectives. | 5-10 |
| **DISCUSSION** | | | |
| Summary of evidence | 19 | Summarize the main results (including an overview of concepts, themes, and types of evidence available), link to the review questions and objectives, and consider the relevance to key groups. | 11-12 |
| Limitations | 20 | Discuss the limitations of the scoping review process. | 12 |
| Conclusions | 21 | Provide a general interpretation of the results with respect to the review questions and objectives, as well as potential implications and/or next steps. | 12 |
| **FUNDING** | | | |
| Funding | 22 | Describe sources of funding for the included sources of evidence, as well as sources of funding for the scoping review. Describe the role of the funders of the scoping review. | 1 |

JBI = Joanna Briggs Institute; PRISMA-ScR = Preferred Reporting Items for Systematic reviews and Meta-Analyses extension for Scoping Reviews.
* Where *sources of evidence* (see second footnote) are compiled from, such as bibliographic databases, social media platforms, and Web sites.
† A more inclusive/heterogeneous term used to account for the different types of evidence or data sources (e.g., quantitative and/or qualitative research, expert opinion, and policy documents) that may be eligible in a scoping review as opposed to only studies. This is not to be confused with *information sources* (see first footnote).
‡ The frameworks by Arksey and O'Malley (6) and Levac and colleagues (7) and the JBI guidance (4, 5) refer to the process of data extraction in a scoping review as data charting.
§ The process of systematically examining research evidence to assess its validity, results, and relevance before using it to inform a decision. This term is used for items 12 and 19 instead of "risk of bias" (which is more applicable to systematic reviews of interventions) to include and acknowledge the various sources of evidence that may be used in a scoping review (e.g., quantitative and/or qualitative research, expert opinion, and policy document).

*From:* Tricco AC, Lillie E, Zarin W, O'Brien KK, Colquhoun H, Levac D, et al. PRISMA Extension for Scoping Reviews (PRISMAScR): Checklist and Explanation. Ann Intern Med. 2018;169:467–473. doi: 10.7326/M18-0850.

### 2.Expanded methods

2.1 Study design

This systematic scoping review was conducted in accordance with the Preferred Reporting Items for Systematic Reviews and Meta-Analyses extension for Scoping Reviews (PRISMA-ScR)[1] and the Joanna Briggs Institute methodology for scoping reviews.[2] The objective of this study aligned with five of the six indications for conducting a scoping review[3] therefore this was the most appropriate method. A strength of scoping reviews is the ability to synthesise evidence from heterogenous disciplines for the purpose of defining concepts and developing evidence-based frameworks, including for vulnerable groups.[4] The protocol was pre-published.[5] An overview of the methods is included below, focusing on areas where the protocol was modified.

2.2 Search strategy

Previous food environment reviews focused on studies that explicitly applied food environment concepts, and synthesised findings deductively using existing food environment frameworks.[6,7] In contrast, we employed wide search criteria to capture all studies reporting on women's food acquisition and dietary intake to inductively identify food environment determinants. A wide criterion was developed based on emerging food environment concepts,[8,9] and the expanded food security definition which includes the new pillars of agency and sustainability.[10] We focused on intermediate food system pathways and outcomes,[11] to unpack the 'missing middle' between food production and nutrition outcomes, aligning with sustainable diet and preventative food-based approaches.[12] By employing this 'wide net' approach, this review aimed to identify all pertinent food environment determinants, especially in low-income countries where explicit food environment research is rare.[6,13]

An academic librarian (MS) guided the selection of databases, and the development and refinement of the search criteria. The search strategy aimed to locate published articles in peer-reviewed journals, utilising a 3-step strategy. An initial limited search of Web of Science Core Collection was undertaken to identify articles on the topic. The text contained in the titles and abstracts of relevant articles were used to develop a full search strategy for Web of Science Core Collection.[5] This search strategy was adapted for each database under the guidance of the academic librarian (MS). The search included 6 databases from the Web of Science Core Collection, and 14 databases from EBSCO and PubMed. In contrast to the protocol, the reference lists of all included sources were not screened manually for additional studies due to the high volume of included studies and resource constraints.

We did not impose limits based on language; however, studies in languages other than English, French, Portuguese, or Spanish were excluded during the screening phase due to resource constraints. Less than 1% of studies were identified in other languages. Due to the evolving nature of food environment determinants in LMICs related to globalisation, urbanisation, and changing agroecological systems, the search strategy was limited to studies published in the last decade between January 1, 2010, and April 30, 2023. The final search was conducted on May 26, 2023.

2.3 Study selection

All identified citations were collated and uploaded into Mendeley V1.19.4 (Mendeley Ltd., Elsevier, Netherlands) and duplicates removed. Screening involved a two-step approach. First, titles and abstracts were screened for assessment against the inclusion criteria using a screening tool on Google Docs that was iteratively developed by the team. The full text of selected citations was then assessed in detail against the inclusion criteria. The study selection process and exclusion reasons were recorded and are reported in the PRISMA flow chart (**Figure 1**). Any disagreements were resolved through group discussion. Each study was assessed in duplicate by members of the team fluent in the relevant language. English studies were assessed by LO, TH, RH and JdB while studies published in Spanish, Portuguese and French were assessed by MFP and PDS. Three final year undergraduate nutrition and public health students also assisted with early stages of title and abstract screening.

From 3,751 retrieved studies, 2,929 were excluded during screening of titles and abstracts. Of the 822 full text articles assessed for eligibility, 304 were excluded with reasons (**Figure 1**). A total of 518 studies were included in the review, with most in English ($n$=502), and fewer in Spanish ($n$=10), Portuguese ($n$=5), and French ($n$=1). The most common reasons for exclusion were as follows: data was not disaggregated by gender, more than 10% of sample size was outside target age range/age was unable to be determined, the study only reported on household or child level outcomes, and the study did not include an outcome variable of interest (e.g., weight status of women was reported but not food acquisition or dietary intake outcomes).

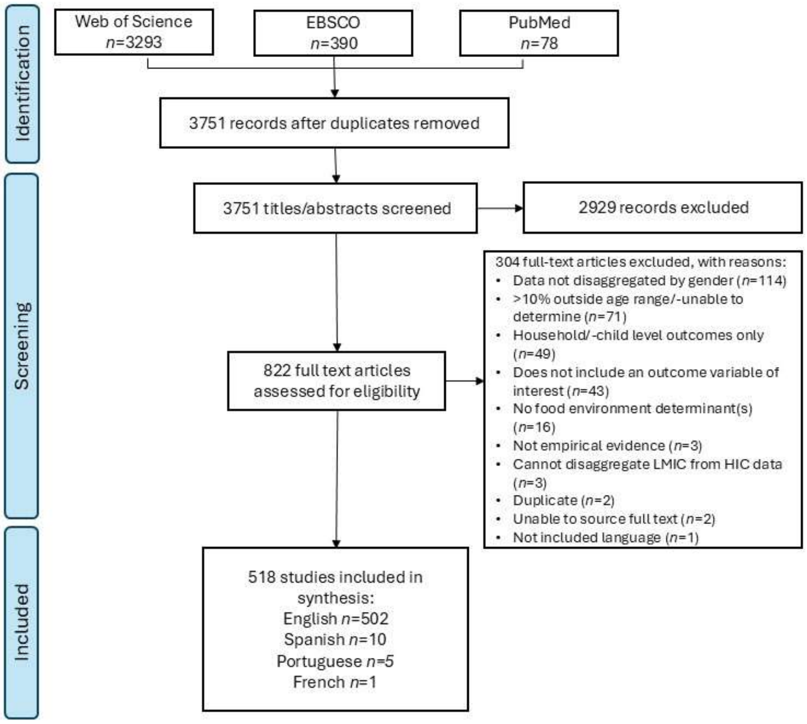

**Figure 1**: Prisma flow chart of included studies. Articles were published between 1 January 2010 and 30 April 2023.

### 2.4 Inclusion and exclusion criteria

The inclusion and exclusion criteria were formulated according to the Participants, Concept, and Context (PCC) tool for scoping reviews. This review included quantitative, qualitative, mixed method, and review studies conducted in any low- and middle-income country (LMIC) as defined by the World Bank for the fiscal year 2021 that described at least one food environment determinant and reported on its influence on at least one food acquisition practice or dietary intake measure for women of reproductive age (WRA). We excluded studies that did not report on outcomes for WRA. If a study also included high-income countries, then it was retained if the data for LMICs was disaggregated. This review included studies of pregnant, lactating, and non-pregnant and non-lactating (NPNL) WRA (15 to 49 years), and participants that identified as the female gender. Quantitative studies that included other genders or ages were retained if the data was disaggregated enabling extraction of data for WRA or if the proportion of the females outside of the target age range was less than 10% of the sample size. For qualitative studies, the study was retained if WRA participated. Gray literature was excluded due to resource constraints.

### 2.5 Data extraction

Data was extracted by LO, TH, RH, JdB, MFP, and PDS using a Google Docs tool iteratively developed by LO and TH in collaboration with the wider team. The data extracted included specific details about the participants, concept, context, study design, methods, and key findings relevant to the review questions. To ensure the comprehensive identification of food environment determinants, drivers of food acquisition and dietary intake were first extracted inductively using an iterative approach. To investigate differences and similarities between physiological stages of heightened nutritional requirements, data was extracted and analysed for pregnant, lactating, and NPNL/unspecified physiological status. Data on food security was extracted if it was a determinant of an outcome of interest; in other words, if food security was only presented as an outcome variable in and of itself, then it was not extracted. Contrary to the protocol, data was extracted singularly due to resource constraints; however, complex studies were discussed as a team to ensure data consistency and quality. Risk of bias assessment of the literature was not conducted because it is not recommended for scoping reviews that aim to provide an evidence map of the breadth and depth of research conducted on a concept.

2.6 Data synthesis and framework development

The patterns of food acquisition and dietary intakes in relation to food environment determinants were charted, mapped, and summarised in tabular and graphical formats. Factors influencing food acquisition practices and dietary intakes were then mapped against dimensions of existing food environment frameworks to identify novel dimensions that may not be represented in current frameworks.[8,9,13–16] Data was then synthesised in a narrative summary that aligned the results to the review questions and highlighted gaps in the literature.

Based on preliminary results, a draft conceptual framework was developed to facilitate informal peer-review with multi-disciplinary colleagues at two international academic conferences in May and June 2023. Preliminary results and the draft conceptual framework were presented by the lead author at the international Agriculture for Nutrition and Health (ANH) Conference in Lillonge, Malawi in June 2023 (ANH23), and with colleagues at the Natural Resources Institute, University of Greenwich at a Food Systems for Nutrition conference in May 2023. The conceptual framework was also reviewed and feedback incorporated from an ANH23 Food Environment side event and discussed with key ANH Food Environment working group members. The selection of final framework determinants was realised in iterative discussion between the authors.

### 3.Descriptive statistics

3.1 Studies by publication year

**Figure 2**: Number of studies by year of publication (*n*=518).

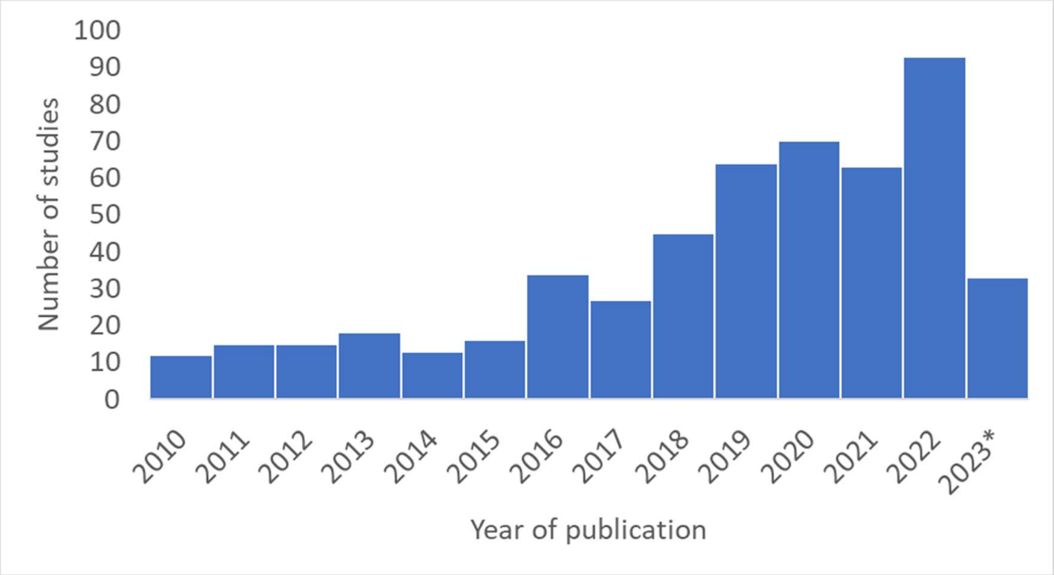

**Includes studies published up to 30 April 2023.*

3.2 Studies by country

**Table 1**: Number of studies by country (*n*=125).

| | Country name | Number of studies |
|---|---|---|
| 1 | Afghanistan | 3 |
| 2 | Albania | 2 |
| 3 | Algeria | 2 |
| 4 | Angola | 3 |
| 5 | Argentina | 4 |
| 6 | Armenia | 1 |
| 7 | America Samoa | 1 |
| 8 | Azerbaijan | 1 |
| 9 | Burundi | 4 |
| 10 | Belarus | 1 |
| 11 | Belize | 1 |
| 12 | Benin | 9 |

| | | |
|---|---|---|
| 13 | Burkina Faso | 17 |
| 14 | Bangladesh | 34 |
| 15 | Bulgaria | 2 |
| 16 | Bhutan | 2 |
| 17 | Bolivia | 4 |
| 18 | Bosnia and Herzegovina | 2 |
| 19 | Brazil | 45 |
| 20 | Botswana | 5 |
| 21 | Cabo Verde | 4 |
| 22 | Central African Republic | 4 |
| 23 | Chad | 2 |
| 24 | China | 20 |
| 25 | Cameroon | 11 |
| 26 | Congo, Dem. Rep. | 9 |
| 27 | Colombia | 11 |
| 28 | Comoros | 2 |
| 29 | Congo, Rep. | 2 |
| 30 | Costa Rica | 6 |
| 31 | Côte d'Ivoire | 2 |
| 32 | Cuba | 1 |
| 33 | Djibouti | 1 |
| 34 | Dominica | 1 |
| 35 | Dominican Republic | 1 |
| 36 | Ecuador | 7 |
| 37 | Egypt, Arab Rep. | 10 |
| 38 | El Salvador | 2 |
| 39 | Equatorial Guinea | 2 |
| 40 | Eritrea | 1 |
| 41 | Ethiopia | 59 |
| 42 | Fiji | 3 |
| 43 | Gabon | 2 |
| 44 | Gambia, The | 4 |
| 45 | Ghana | 18 |
| 46 | Guinea | 6 |
| 47 | Grenada | 1 |
| 48 | Guatemala | 9 |
| 49 | Guinea-Bissau | 3 |
| 50 | Haiti | 1 |
| 51 | Honduras | 3 |
| 52 | Indonesia | 16 |
| 53 | India | 68 |
| 54 | Iraq | 2 |
| 55 | Iran, Islamic Rep. | 16 |
| 56 | Jamaica | 2 |
| 57 | Jordan | 5 |
| 58 | Kazakhstan | 2 |
| 59 | Kenya | 40 |
| 60 | Cambodia | 12 |
| 61 | Kyrgyz Republic | 1 |
| 62 | Lao PDR | 8 |
| 63 | Lebanon | 6 |
| 64 | Libya | 2 |
| 65 | Lesotho | 2 |
| 66 | Liberia | 3 |
| 67 | Sri Lanka | 9 |
| 68 | Maldives | 3 |
| 69 | Morocco | 5 |
| 70 | Mauritania | 3 |
| 71 | Mauritius | 2 |
| 72 | Madagascar | 6 |

| | | |
|---|---|---|
| 73 | Mexico | 16 |
| 74 | Mali | 8 |
| 75 | Myanmar | 6 |
| 76 | Moldova | 1 |
| 77 | Mongolia | 1 |
| 78 | Montenegro | 1 |
| 79 | Mozambique | 12 |
| 80 | Malawi | 22 |
| 81 | Malaysia | 13 |
| 82 | Namibia | 4 |
| 83 | Niger | 5 |
| 84 | Nigeria | 14 |
| 85 | Nicaragua | 2 |
| 86 | North Macedonia | 2 |
| 87 | Nepal | 24 |
| 88 | Pakistan | 15 |
| 89 | Panama | 1 |
| 90 | Papua New Guinea | 1 |
| 91 | Paraguay | 1 |
| 92 | Peru | 7 |
| 93 | Philippines | 9 |
| 94 | Romania | 1 |
| 95 | Russian Federation | 2 |
| 96 | Rwanda | 5 |
| 97 | Samoa | 1 |
| 98 | São Tomé and Principe | 2 |
| 99 | Senegal | 5 |
| 100 | Serbia | 1 |
| 101 | Solomon Islands | 2 |
| 102 | Sierra Leone | 3 |
| 103 | Somalia | 3 |
| 104 | South Sudan | 2 |
| 105 | St Lucia | 2 |
| 106 | St Vincent and the Grenadines | 1 |
| 107 | Sudan | 3 |
| 108 | Suriname | 1 |
| 109 | Eswatini | 3 |
| 110 | Syrian Arab Republic | 1 |
| 111 | Tajikistan | 2 |
| 112 | Thailand | 11 |
| 113 | Timor-Leste | 2 |
| 114 | Togo | 2 |
| 115 | Tonga | 1 |
| 116 | Tunisia | 4 |
| 117 | Turkey | 6 |
| 118 | Tanzania | 34 |
| 119 | Uganda | 19 |
| 120 | Ukraine | 2 |
| 121 | Vietnam | 13 |
| 122 | Yemen, Rep. | 2 |
| 123 | South Africa | 23 |
| 124 | Zambia | 7 |
| 125 | Zimbabwe | 4 |

Note: will not sum to 518 because some studies covered multiple countries. Some of the multi-country studies were not included in above calculations because the countries could not be identified from the text.[17–22]

3.3 Determinants by frequency and region

**Table 2**: Food environment determinants influencing the food acquisition practices and dietary intakes of women in LMICs, by key themes (*n*=15) and frequency for all included studies (*n*=518) and by region.

| Overarching theme | | All (*n*=518) | | Sub-Saharan Africa (*n*=213) | | South Asia (*n*=107) | | Latin America & Caribbean (*n*=81) | | East Asia & Pacific (*n*=62) | | Middle East/North Africa (*n*=28) | | Multi-region (*n*=21) | | Europe & Central Asia (*n*=6) | |
|---|---|---|---|---|---|---|---|---|---|---|---|---|---|---|---|---|---|
| | | ***n*** | **%** | ***n*** | **%** | ***n*** | **%** | ***n*** | **%** | ***n*** | **%** | ***n*** | **%** | ***n*** | **%** | ***n*** | **%** |
| **1** | **Women's agency** | 186 | 35.9 | 88 | 41.3 | 52 | 48.6 | 18 | 22.2 | 9 | 14.5 | 7 | 25.0 | 12 | 57.1 | 0 | 0.0 |
| **2** | **Stability and sustainability** | 169 | 32.6 | 91 | 42.7 | 36 | 33.6 | 17 | 21.0 | 13 | 21.0 | 2 | 7.1 | 8 | 38.1 | 2 | 33.3 |
| **3** | **Food security** | 117 | 22.6 | 55 | 25.8 | 24 | 22.4 | 13 | 16.0 | 13 | 21.0 | 7 | 25.0 | 4 | 19.0 | 1 | 16.7 |
| **External Food Environment (national/regional/institutional/community levels)** | | | | | | | | | | | | | | | | | |
| **4** | **Food availability** | 258 | 49.8 | 126 | 59.2 | 58 | 54.2 | 30 | 37.0 | 26 | 41.9 | 2 | 7.1 | 14 | 66.7 | 2 | 33.3 |
| **5** | **Vendor or food properties** | 130 | 25.1 | 42 | 19.7 | 23 | 21.5 | 30 | 37.0 | 17 | 27.4 | 8 | 28.6 | 9 | 42.9 | 1 | 16.7 |
| **6** | **Food prices** | 102 | 19.7 | 38 | 17.8 | 15 | 14.0 | 21 | 25.9 | 15 | 24.2 | 2 | 7.1 | 9 | 42.9 | 2 | 33.3 |
| **7** | **Food marketing, policies and regulation** | 66 | 12.7 | 16 | 7.5 | 14 | 13.1 | 14 | 17.3 | 10 | 16.1 | 3 | 10.7 | 9 | 42.9 | 0 | 0.0 |
| **Personal Food Environment (social/household/individual levels)** | | | | | | | | | | | | | | | | | |
| **8** | **Food literacy** | 296 | 57.1 | 121 | 56.8 | 66 | 61.7 | 37 | 45.7 | 36 | 58.1 | 24 | 85.7 | 10 | 47.6 | 2 | 33.3 |
| **9** | **Affordability** | 274 | 52.9 | 114 | 53.5 | 49 | 45.8 | 39 | 48.1 | 37 | 59.7 | 18 | 64.3 | 14 | 66.7 | 3 | 50.0 |
| **10** | **Accessibility** | 203 | 39.2 | 83 | 39.0 | 40 | 37.4 | 27 | 33.3 | 32 | 51.6 | 6 | 21.4 | 15 | 71.4 | 0 | 0.0 |
| **11** | **Food preferences** | 202 | 39.0 | 75 | 35.2 | 44 | 41.1 | 36 | 44.4 | 30 | 48.4 | 8 | 28.6 | 7 | 33.3 | 2 | 33.3 |
| **12** | **Social norms** | 135 | 26.1 | 53 | 24.9 | 37 | 34.6 | 20 | 24.7 | 16 | 25.8 | 1 | 3.6 | 7 | 33.3 | 1 | 16.7 |
| **13** | **Social capital** | 106 | 20.5 | 44 | 20.7 | 24 | 22.4 | 16 | 19.8 | 11 | 17.7 | 3 | 10.7 | 7 | 33.3 | 1 | 16.7 |
| **14** | **Convenience** | 86 | 16.6 | 28 | 13.1 | 21 | 19.6 | 16 | 19.8 | 13 | 21.0 | 0 | 0.0 | 7 | 33.3 | 1 | 16.7 |
| **15** | **Food safety perspectives** | 36 | 6.9 | 14 | 6.6 | 5 | 4.7 | 5 | 6.2 | 8 | 12.9 | 1 | 3.6 | 3 | 14.3 | 0 | 0.0 |

### 4.Glossary of terms

**Table 3:** Glossary of terms of key food environment determinants included in the conceptual framework and findings of this study.

| Term | Definition | Reference |
|---|---|---|
| **Accessibility** | Physical access to food. Accessibility is relative to individuals. Accessibility is highly dynamic and can include distance, time, space and place, daily mobility, and modes of transport that collectively shape individual activity spaces.<br>See also ***Availability***, and ***Freedom of movement***. | [8] |
| **Affordability** | Economic access to food. Affordability is determined by the interaction between food prices and household and/or individual purchasing power. Affordability is mediated by an individuals' financial autonomy and agency.<br>See also ***Prices***. | [8] |
| **Agency** | Agency refers to the capacity and power of an individual to exercise control over their own circumstances and to provide meaningful input into governance processes. In food environments, it refers to the capacity or power of an Individual to act independently to make choices and exert control over food, including about the foods they acquire and consume. The protection of individual agency requires socio-political systems that uphold governance structures that enable decision-making autonomy, bargaining power, financial autonomy, control over time, and freedom of movement to enable an individual to exert control over resources for their own dietary and nutritional needs.<br>See also ***Bargaining power, Power imbalance, Coercive control, Decision-making autonomy, Financial autonomy, Control over time, and Women's empowerment.*** | Adapted from [10,23–25] |
| **Availability** | Physical availability of food. Availability refers to whether or not a vendor or product is present within a given context. Availability always precedes accessibility (i.e., a food cannot be accessible if it is not available). Food sources in LMICs include cultivated, informal and formal markets, wild harvest, food aid (including national feeding programmes) and social exchange (e.g., gifts, remittances, barter).<br>See also ***Accessibility***. | Adapted from [8] |
| **Bargaining power** | An individuals' agency (power) to negotiate favourable allocation of resources at societal and intra-household levels. An individual's level of education, income and assets are important aspects of bargaining power. Regarding nutritional outcomes of food environments, an individual's bargaining power may affect intra-household allocation of resources related to the procurement and consumption of food, and aspects of household production or time use, including the allocation of labour to various activities (paid and unpaid). The outside options of individuals affect their bargaining power within the household. For example, increases in an individual's income, assets, and land or divorce rights may give an individual more bargaining power within a household or marriage. Bargaining power may also affect individual well-being by reducing risk of elements of coercive control | Adapted from [26] |

| | | |
|---|---|---|
| | and/or gender-based violence affecting food acquisition and dietary intakes. See also ***Agency, Power imbalance, Coercive control, Decision-making autonomy, Financial autonomy, Control over time, and Women's empowerment.*** | |
| **Coercive control** | Course of behaviours aimed at dominating and controlling another by diminishing the ability of an individual to exercise agency and autonomy. Coercive control is a manifestation of power, where a perpetrator uses non-physical (e.g. emotional or financial abuse) and/or physical tactics (violence, or threat thereof) to make another subordinate and maintain dominance. Coercive control is often of an ongoing, repetitive and cumulative nature. The attack on an individual's autonomy can involve strategies such as physical, sexual, verbal and/or emotional abuse, psychologically controlling acts, and deprivation of resources. In relation to dietary and nutrition outcomes, coercive control behaviours such as financial abuse, withholding of food, and constraints on freedom of movement can affect what foods are procured or consumed. In coercive or violent environments, an individual may demonstrate 'burdened agency' wherein they allocate household resources (e.g., food or money) to the perpetrator in the hopes of preventing violence or abuse. See also ***Agency, Power imbalance, Decision-making autonomy***, and **Bargaining power**. | Adapted from [27,28] |
| **Control over time** | The autonomy and ability to make and act upon strategic choices about how to allocate one's time. The concept of control over time (and time use agency) links empowerment and time use by shifting focus away from how much time one spends on certain activities toward the agency (power of) an individual to make strategic choices about how to allocate their own time. See also ***Time use, Time poverty, Agency, Decision-making autonomy, and Women's empowerment***. | Adapted from [29] |
| **Convenience** | A characteristic that results in reduced requirement for resources including time, physical effort, mental effort and skills by the consumer in relation to the planning, acquisition, preparation, storage, transport, consumption or clean-up of food. See also ***Time use, Control over time, Time poverty, and Accessibility.*** | [30] |
| **Decision-making autonomy** | The idea of decision-making autonomy emphasises the individual's rights to self-govern and make choices without interference or control by others. In the context of food choices, decision-making autonomy relates to the autonomy of the individual to make decisions about what foods (and how much) to grow, acquire/purchase, keep for home consumption, and eat. The degree of decision-making control that an individual experiences can occur along a spectrum, differing by eating location, occasion, and social context. See also ***Agency, Financial autonomy***, ***Freedom of movement***, **Bargaining power**, and ***Social norms***. | Adapted from [23,31,32] |
| **Financial autonomy** | The concept of financial autonomy is the ability of an individual to make autonomous decisions related to their income, expenses, and level of debt. In the context of food choices, financial autonomy relates to the autonomy of the individual to make decisions about how to earn/procure | Adapted from [23,31] |

| | | |
|---|---|---|
| | money and how to spend money to acquire food (e.g., decision-making autonomy on how to spend money on transport costs to access food sources, and food purchases).<br>See also ***Agency, Bargaining power, Freedom of movement,*** and ***Social norms***. | |
| **Food acquisition** | The act of acquiring food by individuals and/or households, including through purchases and other transactions, production, harvesting and gifts. Food acquisition practices may be considered as a key outcome of a given food environment, separate to food consumption.<br>See also **Non-transactional food acquisition**, and **Food purchasing** | Authors' own |
| **Food literacy** | A collection of inter-related knowledge, skills and behaviours required to make informed decisions and actions related to planning, managing, selecting, preparing and consuming foods to meet needs and determine intake. Food literacy extends beyond knowledge to also include practical skills relating to food procurement, storage, and preparation.<br>See also ***Food preferences***. | Adapted from [33] |
| **Food security** | Food security is a situation that exists when all people, at all times, have physical, social and economic access to sufficient, safe and nutritious food that meets dietary needs and food preferences for an active and healthy life. | [10,34,35] |
| **Food preferences** | The evaluative attitudes that people express towards preferred or desired foods or diets, shaped by sensory aspects (e.g. taste, smell, texture, mouth-feel), morbidity (e.g., disease status) or physiological status (e.g. pregnancy, lactation), cultural (e.g. food taboos, 'prestige' foods), religious (e.g. food restrictions or promotions), and personal aspirations (e.g. vegetarian).<br>See also ***Food literacy.*** | [8,33] |
| **Food properties** | Food properties refer to aspects such as quality, safety, level of processing, shelf-life and packaging.<br>See also ***Vendor properties***. | Adapted from [8] |
| **Food purchasing** | Transactional food acquisition (e.g., buy foods from markets / exchange goods or services for food).<br>See also ***Non-transactional food acquisition***. | Authors own |
| **Food safety perspectives** | An individual's perspectives about agrochemicals, adulteration, food hygiene or environmental sanitation, or livestock health.<br>See also ***Food properties, and Vendor properties***. | Adapted from [8,36] |
| **Food sovereignty** | The right of individuals to healthy and culturally appropriate food produced through socially just and ecologically sensitive methods. It entails people's right to participate in food system and food environment decision-making and define their own food, agriculture, livestock and fisheries systems, and food environment food sources (market and non-market).<br>See also ***Sustainability***. | Adapted from [37] |
| **Freedom of movement** | Freedom of movement can be shaped by the socio-cultural decision-making autonomy of an individual to choose to leave the house by themselves, how or if an individual feels safe walking or catching public transport to procure food from food sources, and if they have the financial autonomy to afford and pay for transport fees. | Adapted from [31,38] |

| | | |
|---|---|---|
| | See also ***Agency, Coercive control, Decision-making autonomy, Bargaining power, Financial autonomy***, ***Social norms***, and ***Accessibility***. | |
| **Marketing** | Marketing includes promotional information, branding, advertising, sponsorship, and labelling pertaining to the sale of foods.<br>See also ***Regulation***. | Adapted from [8,34] |
| **Non-transactional food acquisition** | Non-transactional food acquisition (e.g., harvest home production, gather wild foods).<br>See also ***Food acquisition, and Food purchasing.*** | Authors own |
| **Power imbalance** | A position in which both or all the groups or people involved do not have equal power. In practice, it occurs when a partner or group can dominate decision making or otherwise asserts power in ways that disadvantage other partners or is not in their best interest.<br>See also ***Coercive control, Agency***, ***Decision-making autonomy,*** *and* ***Bargaining Power***. | Adapted from [27,28] |
| **Prices** | Prices refer to the cost of food products. Prices interact with individual and household purchasing power to determine affordability. Prices and affordability are sensitive to fluctuations in food availability and accessibility.<br>See also ***Affordability***. | Adapted from [8] |
| **Regulation and policies** | Regulation includes policies pertaining to production, distribution, and sale of foods.<br>For example, regulations can include providing incentives for improving the nutritional quality of processed foods and their promotion in food retail and advertising, as well as disincentives for non-adherence. Regulations or policies can also pertain to incentives or disincentives relating to food production, including the cultivated foods kept for home consumption.<br>See also ***Marketing***. | Adapted from [8,34] |
| **Social capital** | The sum of the actual and potential resources embedded within, available through, and derived from the network of relationships possessed by an individual or social unit. Social capital thus comprises both the network and the resources that may be mobilised through that network. The characteristics of a social network determine the social capital of its individual actors. In contrast to physical and human capital, social capital focuses on relations between people.<br>See also **Social norms, and Agency**. | Adapted from [39,40] |
| **Socio-cultural behaviours/practices** | See ***Social norms.*** | |
| **Social networks** | See ***Social capital***. | |
| **Social norms** | Social norms are the perceived informal, spoken or unspoken rules, about what behaviors are appropriate, typical, or obligatory within a given group and reflect the influence of community and household beliefs on individual behavior.<br>Socio-cultural norms, alongside the influence of kin and community, households, spousal, children, and peers' food preferences and habits, can influence individual decision-making about food procurement, preparation, communal sharing, and consumption. Norms can also shape socio-cultural expectations of what is appropriate or typical | Adapted from [41,42] |

| | | |
|---|---|---|
| | regarding an individual's access to resources, including food and time for nutrition-related activities.<br>See also ***Agency, Social capital***, ***Food literacy***, and ***Food preferences***. | |
| **Stability** | Having the ability to ensure sufficient food acquisition and consumption in the event of sudden shocks (e.g., an economic, health, conflict or climate crisis) or cyclical events (e.g., seasonal food insecurity). For example, seasonal fluctuations in production and price of foods, limited reliable availability and affordability of fruits and vegetables, variations in climate, seed quality and availability, pests can harm crop production, and income variability affected choices about food purchasing due to unreliable affordability of foods.<br>See also ***Sustainability***. | [10,34] |
| **Sustainability** | Food system and food environment practices that contribute to long-term regeneration of natural (environmental), social, and economic systems, ensuring the food needs of the present generations are met without compromising the food needs of future generations.<br>See also ***Stability***. | [10,34] |
| **Time use** | The influence of an individual's time allocation on the decision-making process of food acquisition, preparation, and consumption. For example, an individual's workloads (paid and unpaid) and livelihoods outside of the home may impact time available for food acquisition and preparation, altering food choices for the individual and – in strict gender norm settings - the whole household.<br>See also ***Control over time, and Time poverty***. | Adapted from [29,43] |
| **Time poverty** | A state of deprivation characterised by the reduced ability to make unconstrained choices about one's personal time allocation brought about by working long hours.<br>See also ***Control over time, and Time use***. | [43] |
| **Vendor properties** | Vendor properties refer to aspects such as the type of food vendors, opening hours, and services provided.<br>See also ***Food properties***. | [8] |
| **Women's empowerment** | Process by which women gain power and control over their own lives and acquire the ability to make strategic choices. Empowerment is "a process of the expansion of the ability to make strategic life choices in a context where this ability was previously denied". Women's empowerment implies freedom to make choices around food acquisition, production, and consumption, which may affect dietary quality. In some contexts, this capacity may be countered by other household members, such as male household heads, husbands, older women (e.g., mother-in-law, aunt).<br>See also ***Agency, Decision-making autonomy, Financial autonomy, Power imbalance***, and ***Coercive control.*** | Adapted from [23–25,31] |

**5.Determinant themes of food acquisition and dietary intakes**

**Table 4**: Food environment determinants that influence the food acquisition practices and dietary intakes of women in LMICs, by overarching themes (*n*=15), secondary (*n*=30), and tertiary levels (*n*=143) and corresponding references.

| | Theme – level 1 (*n*=15) | Theme – level 2 (*n*=30) | Theme – level 3 (*n*=143) | References |
|---|---|---|---|---|
| **1** | **(1) Women's agency** | **Decision-making autonomy and bargaining power** | Decision making autonomy and bargaining power to influence **food procurement, preparation and consumption decisions** (i.e., food choice) | 21,44–86 |
| **2** | | | Decision making autonomy and bargaining power to influence **food production decisions** | 44,54,57,60,65,72,87–98 |
| **3** | | | Decision making autonomy and bargaining power to influence **decisions about home consumption of produced foods** | 20,44,60,65,66,69,77,87–89,93–105 |
| **4** | | | Lack of decision-making autonomy and bargaining power around food influenced by **intra-household power imbalances** due to **socio-cultural norms privileging decision-making power and access to and control over resources to other household members** such as male head of household (e.g., husband) or older women (e.g., mother-in-law) | 20,44,45,47,50,53,54,56,59–61,63–65,67–69,71,74,77,78,83,84,94,95,100,101,106–117 |
| **5** | | | Denial of food and dependence on others for food acquisition and allocation due to **violations of decision-making and financial autonomy around food and freedom of movement to procure food** enforced by **coercive control and gender-based violence**, including manipulation or other controlling behaviour(s) | 45,47,54,63–65,69,73,74,77,81,94,97,100,101,106,107,113,114,118,119 |
| **6** | | | **Lack of decision-making autonomy and bargaining power regarding intra-household allocation of food** influenced by **socio-cultural norms** such as feeding other members of the household first (e.g., men, elders, children/maternal buffering) and other factors | 17,19–21,45,47,50,56,57,59–61,64,65,67–71,73–76,78,79,81,87,93,97,102,106,108–110,113,114,118,120–138 |

| | | | | |
|---|---|---|---|---|
| **7** | | | Lack of decision-making autonomy and bargaining power regarding **intra-household allocation of food** influenced by **household size and number of household dependents** (e.g., 'mouths to feed'), affecting the types and amounts of foods available for women to eat (e.g., household food insecurity exacerbating negative effects of socio-cultural norms driving intra-household food allocation, especially if women served last). | 19,47,55,56,62–65,79,85,87,93,99,108,113,115,118,120,122,123,132,136–180 |
| **8** | | | Decision-making autonomy and bargaining power related to **women's empowerment** (e.g., women's empowerment in agriculture/-livestock indexes, Demographic and Health Survey empowerment indicators) | 20,62,63,65,84,90,93,98,100,101,108,181–185 |
| **9** | | **Financial autonomy** | **Women's financial independence and control over income** | 20,22,48,52,58–60,62,64–66,68,69,76–78,81,84,88,91,93–95,97,100,101,103,106,108,109,111–113,115–117,146,156,165,170,173,181,186–198 |
| **10** | | | Autonomy to make **financial decisions about food** | 20,21,44,45,48,49,55,57–60,63,65,66,68,69,73,74,76–78,81,84,87,91,93–95,100,108,111,113,116,147,156,170,191,199,200 |
| **11** | | | **Women's livelihoods outside of the home** | 20,49,58,63,66,69,76,77,86,88,97,106,109,111,112,119,120,138,156,159,165,185,190,194–196,201–204 |
| **12** | | **Control over time** | **Women's workloads and control over time** | 19,20,22,60,61,64,65,68,69,71,79,81,82,86,88,91,92,94–98,101,102,111,117,138,156,185,188,190,192–194,198,202,205–218 |
| **13** | | | **Sociocultural gender norms** (e.g., women identified as food providers which increases their unpaid caregiving duties, influencing workloads, time poverty, and control over time) | 17,19,20,44,47,49–52,59–61,64,65,70–72,76–79,81,82,88,91–95,97,107,111,112,114,117,133,188,191,202,211,214,215,219 |
| **14** | | **Freedom of movement** | **Freedom of movement** including physical safety (e.g. fear of violence) and **socio-cultural norms** influencing autonomy to leave the house and mobility to procure food | 20,64,65,69,70,73,74,81,94,95,99,106,111,117,118,120,156,181,193,194,212 |
| **15** | **(2) Stability and sustainability** | **Stability** | **Seasonal fluctuations in food availability** | 19,20,22,44,47,48,51,52,54,57,59,60,64,68,69,71,75,77,87,88,92,94,96,97,99,101–103,109,110,113,118,132,133,136,144–146,148,149,154,155,161,165,174,179,186,191,196,197,211,212,217,220–269 |
| **16** | | | **Seasonal fluctuations in food prices** | 44,51,59,75,77,88,89,94,103,113,133,136,211,212,223,249,254,257,260,270,271 |
| **17** | | **Variations in food availability/ food shortages** due to… | **Instability of purchasing power, including due to income variability / precarious livelihoods / daily survival / difficulties accessing social safety nets** (e.g. registration / payment delays) | 19,20,58,61,64,70,77,86,88,94,105,112,113,133,136,138,140,162,165,188,195–197,202,240,250,257–260,272–274 |
| **18** | | | **Drivers such as climate change** | 54,60,64,77,92,94,102,113,136,138,148,149,186,196,212,220,223,231,237,245,254,266,268 |

| | | | | |
|---|---|---|---|---|
| **19** | | | **Acute shocks (e.g., Covid-19 pandemic, epidemics, conflicts, floods, droughts)** | 20,62,92,111,119,120,126,135,156,194,198,202,212,227,252,254,268,273,275–281 |
| **20** | | | **Water availability** | 68,144,196 |
| **21** | | | **Transhumance/nomadic practices** | 94,101,268 |
| **22** | | | **Globalization and trade liberalisation** | 88,282 |
| **23** | | **Sustainability** | **Agrobiodiversity** | 57,91,152,154,165,221,237,248,258,266,283–290 |
| **24** | | | **Indigenous foods** | 57,94,131,234–237,270,271,285,287,291–294 |
| **25** | | | **Sustainable diets / Carbon footprint of food** | 19,195,295–299 |
| **26** | | | **Agroecological farming practices / climate adaptation** | 54,92,94,286,290 |
| **27** | | | **Loss of agrobiodiversity / natural resource base** | 57,94,101,138,231,266,300,301 |
| **28** | | | **Culturally appropriate food supply / food sovereignty** | 46,54,57,88,94,200,231,237,263,296,302 |
| **29** | | | **Traditional food knowledge** | 57,92,165 |
| **30** | **(3) Food security** | **Food security** | **Household food security** (e.g., Household Food Insecurity Access Scale, Food Consumption Score, household food expenditure) | 17,56,58,62,63,80,87,88,97,109,118,120,123,132,137,138,141,150–153,159,160,162,165,168,173–175,180,198,220,228,233,240,244,251,252,256,267,276,282,284,303–323 |
| **31** | | | **Individual food security** (e.g., Individual Food Insecurity Experience Scale) | 101,106,140,149,169,187,206,220,324–327 |
| **32** | | **Food insecurity behaviours / perceived concerns** | Moderate to severe food insecurity coping behaviours, such as **meal skipping/-rationing food/-limiting food intake** | 47,49,61,62,64,69–71,75,79,86,99,102,103,111,113,119,120,122,126,133,138,176,188,194,197,202,206,212,250,253,262,269,272,273,304,310,328–331 |
| **33** | | | Respondent reported **"hungry all the time" / "worried about where the next meal is coming from"** | 69,70,102,112,113,120,122,133,194,206,212,240,250,272,304,310,331 |
| **EXTERNAL FOOD ENVIRONMENT (NATIONAL, REGIONAL, INSTITUTIONAL, AND COMMUNITY LEVELS)** | | | | |
| **34** | **(4) Food availability** | **Food source** | Food available from **home production** (includes crops, livestock and land ownership) | 19,20,22,44,48,51,52,54,55,57,60,63,65,66,68,69,71,75–77,80,84,87,88,91–105,114,115,118,121,124,127,132,133,136,138,144–148,152,154,155,165,166,175,177,183,186–189,191,193,194,196,202,206,217,221,223,228,230,231,234,235,237,240–242,246,248,250–255,257–260,264,266,267,269,272,275,276,283,286–290,300,301,306–308,311,324,325,328,329,332–362 |
| **35** | | | Food available from **Markets (general/-unspecified)** | 19,22,46,48,64,65,68,70,76,77,87,88,91,93,94,99,112–114,124,126,127,136,138,145,154,156,166,183,190,191,194,197,200,202,207,211,212,217,236,237,247,249,257,260,269,271,276,277,282–284,290,301,308,328,331,349,353,363–370 |
| **36** | | | Food available from **Informal markets** | 44,49,59,68,70,71,75,82,86,97,115,116,118,122,133,146,148,179,188,211,221,228,233,241,250,254,255,257,259,275,290,300,302,307,309–311,342,358,367,371–376 |

| | | | | |
|---|---|---|---|---|
| **37** | | | Food available from **Formal markets** | 51,52,54,57,82,103,118,122,126,133,146,179,211,218,224,228,233,236,246,254,266,275,290,302,307,309–311,342,367,372,377–381 |
| **38** | | | Food available from the **Wild (e.g. forests, lakes, rivers, oceans)** | 19,51,57,71,87,92,93,99,122,138,146,188,194,195,205,210,217,228,231,234–237,246,248,258,259,266,270,272,276,283,287,291,293,294,300,342,382 |
| **39** | | | Food available from **Food aid (acute response) and national feeding programmes (longer-term)** | 21,22,57,79,80,110,119,120,122,123,126,128,132,135,136,140,157,180,198,216,226,236,237,249,268,272,273,276,277,283,304,311,315,325,326,329,352,383 |
| **40** | | | Food available from **Kin and community** | 54,70,86,92,99,113,120,122,138,194,202,215,228,248,272,275,276,290,307 |
| **41** | | | Food available from **Prepared food outlets** | 82,115,179,188,190,202,211,214,215,257,296,299,328,372,379,384,385 |
| **42** | | | Food available from **Dumpster / discarded food** | 113,122 |
| **43** | | | Food available from **School, university, workplace, refugee camp/-settlement, prison** | 179,215,296,310,349,374,385–388 |
| **44** | | **Markets / food retail outlets** | **Presence/count/density of markets / food retail outlets** | 99,122,146,250,302,308–310,371,377,379,381 |
| **45** | | **Food diversity** | **Farm/agricultural/production diversity/ nutritional functional diversity** | 22,54,57,87,89,92,94,95,124,136,146,147,154,155,182,183,185,186,196,206,220,221,223,230,231,235,236,247,250,255,258,266,269,288,290,300,306,325,335,343,346,353,354,357,368,389 |
| **46** | | | **Wild food diversity** | 57,231,235,236,247,248,258,266,270,287,289,294,300,390,391 |
| **47** | | | **Agrobiodiversity** (i.e., both farm and wild food diversity) | 91,152,165,237,248,258,266,283–287,289 |
| **48** | | | **National food supply / trade** | 88,120,186,194,202 |
| **49** | | | **Foods available in the home** | 82,192,374,392 |
| **50** | **(5) Vendor and food properties** | **Vendor properties** | **Vendor / food source type** | 52,54,57,59,86,120,122,123,126,128,129,133,134,138,154,188,194,199,202,207,211,212,221,224,241,254,257,276,290,299,302,309,310,315,328,342,364,367,369,371,373,376,377,379,381,384,385,393–395 |
| **51** | | **Vendor services** | **Vendor services** (including food delivery options / eCommence / electronic biometric scanning for eVouchers etc) | 59,99,112,194,199,202,245,249,254,257,279,302,310,385 |
| **52** | | | **Vendor opening hours** | 120,135,156,194,202,212,257,275,276 |
| **53** | | | **Vendor offers credit** | 49,59,86,112,120,138,188,207,211,254,257,277,376,379 |
| **54** | | **Food properties** | **Food / product quality** | 17,21,51,59,70,86,113,129,133,195,197,199,202,211,212,219,233,254,257,302,342,362,376,396–399 |
| **55** | | | **Food safety** | 19,52,59,86,113,188,195,199,211,212,217,241,254,257,302,307,342,373,374,376,379,399–403 |
| **56** | | | **Processing** | 88,140,197,199,217,218,241,246,254,257,271,279,280,362,364,385,400,404–410 |
| **57** | | | **Food composition** (e.g., level of fortification, level of macro or micronutrients/-salt/-sugar) | 21,22,79,127–129,132,157,283,367,411–423 |
| **58** | | | **Product labelling** (e.g., front of packet labelling, health claims, country of origin, where food is procured from/how it is produced) | 17,130,199,217,298,367,396,411,424–427 |
| **59** | | | **Packaging** | 17,82,128,199,218,257,367,396,397 |

| | | | | |
|---|---|---|---|---|
| **60** | | | **Perishability / shelf life** | 97,99,120,128,197,233,254,282 |
| **61** | **(6) Food prices** | **Food prices and cost of diet** | **Food prices** | 17,19,22,46,49,51,52,59,62,67,70,75–77,82,88,93,94,103,113,116,120,122,124,126,129,130,133,136,138,156,179,188,193,194,197,199,202,211,212,215,217,233,234,245,249,254,257,260,269,271,272,275–277,279,282,296–299,302,307,309,330,342,349,364,365,369,370,373,374,377–379,390,397,398,403,428–431 |
| **62** | | | **Cost of diet** | 270,271,294,297,415,432–434 |
| **63** | **(7) Marketing, policies, and regulation** | **Marketing** | **Promotional information (general marketing)** | 57,117,179,193,207,224,241,254,257,366,374,376,379,428 |
| **64** | | | **Promotional information**, including through product labels and other messaging (e.g,. health claims, source, sustainability) | 199,211,217,257,364,367,379,411,425,426 |
| **65** | | | **Brand advertising** | 17,82,199,211,217,257,298,379,397,435,436 |
| **66** | | **Policies and regulations** | **Food policies** (e.g., fruit and vegetable tax subsidies, staple food subsidies, control on food import/-export, government food aid and national feeding programmes) | 21,22,88,157,275,302,376,436 |
| **67** | | | **Food policies related to external shocks and disaster response** (e.g. Covid-19 pandemic, cyclones, earthquakes, droughts, famine) | 22,111,120,126,156,176,194,202,227,275–280,401,437 |
| **68** | | | **Food regulation** (e.g. mandatory fortification, food safety) | 22,79,127,132,157,283,302,412,416,417,420,423,438,439 |
| **PERSONAL FOOD ENVIRONMENT (SOCIAL, HOUSEHOLD, AND INDIVIDUAL LEVELS)** | | | | |
| **69** | **(8) Food literacy** | **Nutrition knowledge** | **Knowledge about what to eat based on age/health/physiological status** | 17,19–22,45,48,51,53,59,61,62,64,65,67,68,70,71,73,75,77,79,80,82,83,85–87,91,94–97,102,107,109–111,113–118,121,124,130,132,133,135,136,140,142,148,152,165,168,173,175,179,189,192,193,195,197,198,202,204,208,212,213,216,217,219,226,229,233,240,241,245,250,253,260,266,269,274,283,286,288,296,301,302,304,306,315,320–322,325,326,329,331,337–340,345,347,349,351,352,354,356,357,360,363,364,369–374,377,379,390,391,393,394,400,401,411,418,428,431,436,437,440–467 |
| **70** | | | **Women's level of education** | 17,19,20,53,55,61,62,64,65,71,84,85,102,106,109,110,118,124,138–143,145,146,148,152,154,155,158,159,163,164,167,168,170–179,186,187,201,203,204,208,215,226,231,239,246,250,253,255,256,267,272,274,278,284,289,304–306,311–313,316,318,320,322,324,334,338,344,349,350,353,358,362,363,370,371,375,384,388–390,393,396,398,400,405,410,431,439,449,452,454,458,460,461,464,465,468–495 |
| **71** | | | **Male household head/husband's level of education / nutrition knowledge** | 19,20,61,64,65,71,115,138,143,148,152,161,163,164,173,213,216,250,274,288,291,311–313,320,334,363,368,389,437,443,459,482 |
| **72** | | | **Older female of household's** (e.g mother-in-law, aunt) **level of education / nutrition knowledge** | 19,20,64,65,71,138,216,288,446,459 |

| | | | | |
|---|---|---|---|---|
| **73** | | **Food-related knowledge and skills** | **Knowledge of how to grow food** (including sustainable farming practices) | 20,48,54,57,87,91,92,94–96,121,136,230,236,250,260,266,283,306,325,334,339–341,347,348,351,352,354,356–358,360 |
| **74** | | | **Cooking/ food preparation skills** | 59,68,72,99,104,121,136,148,179,202,212,219,241,245,283,315,337,352,354,372,395,401,451,496 |
| **75** | | | **Knowledge of where /- how to acquire food** | 54,57,59,68,99,112,148,186,213,236,266,272,337 |
| **76** | | | **Food preservation skills and resources** (e.g. solar drying, fermenting) | 97,103,193,195,212,245 |
| **77** | | | **Knowledge of how to store food** to maintain quality and safety | 54,97,193,212,260,367,401,463 |
| **78** | | | **Labelling – understanding/knowing how to interpret food labels** | 367,370,379,396,426,431 |
| **79** | **(9) Affordability** | **Income and wealth** | **Household income** | 19–21,44,55,61,64,65,67–69,71,76,80,87,88,92,97,103,106,110,111,113,116,118,126,136,138,140,142–144,146,148,149,152,153,155,156,158–160,162,165,169,172,173,177–179,186,188,189,191,196,198,201,212,228,231,240,246,247,250,253,256–258,260,266,272–277,284,288,304,305,308,311–313,318,320,324,334,335,350,368,370,389,405,410,431,432,443,449,452,456,461,464,470,472,477,479,482,483,485–487,489–492,494,497–503 |
| **80** | | | **Household wealth** | 19,44,61–64,84,110,118,134,143,150,152,154,155,163,165–169,173,175,184–187,198,220,228,229,243,246,249,256,260,268,270,286,289,300,301,308,316,329,334,342,345,359,363,384,399,465,468,474,478,486 |
| **81** | | | **Individual income / wealth** | 20–22,58,63,66–68,76,77,86,88,94,97,109,112,119,138,154,164,170,171,173,188,190,192,194–197,202,204,206,212,226,257,374,375,475,476,481,488 |
| **82** | | | **Socioeconomic status/- class** | 17,61,64,86,112,113,138,140,141,145,163,179,239,249,250,253,304,322,324,388,395,449,458,460,494,495,504 |
| **83** | | | **Employment and livelihood status,** including diversity (e.g. non-farm income), stability and occupation type | 63,154,163–165,274,482 |
| **84** | | | **Cash transfers / vouchers** | 110,191,196,240,283,356 |
| **85** | | | **Household expenditure and debt** | 46,48,63,432,491 |
| **86** | | **Purchasing power** | **Income relative to food prices** | 19–22,44,61,70,71,77,79,82,94,97,111–116,119,120,136,138,165,174,179,191,193,194,196–198,200,211,212,217,218,223,224,249,257–260,266,270,273,275–277,283,290,299,302,307,326,330,390,415,432,434,469,473 |
| **87** | | | **Perceived affordability of food/food group** | 49,51,52,67,73–75,93,106,113,120,122,133,202,207,211,234,241,245,254,269,296,310,349,354,364,369,379,429,487,505 |
| **88** | | | **Cost of nutrient-rich foods relative to income** (e.g., fruits and vegetables, animal-source foods) | 47,68,79,97,190,202,212,273,282,330,342,364,366,370 |
| **89** | | | **Cost of other foods relative to income or nutrient-rich foods** (eg, ultra-processed foods, ready-to-eat meals, imported processed foods) | 88,120,194,212,366 |

| | | | | |
|---|---|---|---|---|
| **90** | **(10) Accessibility** | | **Geographical differences** (e.g., agro-ecological zones, region v region / district v district) | 18–20,57,60,64,75,84,118,122,124,125,132,136,143,146,149,151,152,155,185,186,195,217,221–223,229–231,244,246,247,253,255,257,278,284,286,301,308,324,326,332,338,342,348,363,365,372,389,398,412,414,416–418,420,432,438,467,471,475,478,483,484,487,490,500,502,504,506–521 |
| **91** | | | **Rural-urban comparison/continuum** | 18–20,61,84,106,131,138,139,146,177,189,200,217,219,228,233,243,253,278,293,295,324,362,367,375,389,417,420,432,463,465,468,474,478,479,483,490,492,500,502,506,507,509–511,513,516,522–528 |
| **92** | | **Physical access to food source(s)** | Physical access to **food sources (general)** | 44,51,52,57,59,68,82,99,122,174,179,202,205,212,221,224,231,241,257,266,275,276,302,308,310,363,374,377,380–382,453 |
| **93** | | | Physical access to **Informal markets** (e.g., produce markets, roadside food sources) | 94,111,120,124,136,146,166,179,194,202,212,237,250,291,302,310,353,368,371 |
| **94** | | | Physical access to **Formal markets** (e.g., supermarkets) food sources | 52,54,57,68,70,91,103,124,146,179,186,194,202,291,302,372 |
| **95** | | | Physical access to **Cultivated (e.g., farm) food sources** | 54,68,91,92,96,103,186,189,231,241,250,291,354 |
| **96** | | | Physical access to **Wild food sources** (e.g., forests, water bodies) | 52,57,68,87,99,122,194,231,245,291,300 |
| **97** | | | Physical access to **Prepared food outlet food sources** | 179,202,214,215,296,310,371 |
| **98** | | | Physical access to **Food aid/-price controlled food sources** | 21,126,148,237,275,277,281 |
| **99** | | **Physical distance** | **Proximity to food source(s)** | 19,87,119,128,135,136,138,152,195,197,202,210–212,214,241,246,247,250,255,257,260,275,300,302,326,334,342,353,369,381 |
| **100** | | | **Travel time** (e.g., to market) | 21,51,57,66,94,136,138,145,154,166,174,179,193,202,211,212,217,224,257,260,288,302,376,381 |
| **101** | | **Mobility** | **Daily mobility / individual activity space** | 17,19,64,69,70,73,81,119,120,136,156,194,202,210,211,279,381,388 |
| **102** | | | **Transport costs** | 20,51,52,88,99,113,211,212 |
| **103** | | | **Access to transport** | 20,94,113,174,204,277,380 |
| **104** | | **Acute shocks – physical access changes** | **Government-mandated restrictions to movement** (e.g. Covid-19 lockdowns) | 111,120,126,156,194,202,275–277 |
| **105** | **(11) Food preferences** | | **Food preferences / desired foods (general)** | 19,44,51–53,57,64,67,68,74,77,78,82,86,92–94,96,97,112–114,116,121,128–130,136,138,165,174,176,179,191,192,197,199,200,202,207,212,214,217–219,224,233,241,254,257,258,260,262,268,272,280,282,283,296,298,301,309,326,328,331,342,349,358,362,366,369,372,374,377,379,385,394,395,398,400,424,426,431,444,455,460,463,464,473,498,512,518,529–532 |
| **106** | | | **Perceived healthfulness of food** | 45,50–53,67,68,70,77,82,87,96,114,116,129,133,136,148,157,188,192,197,202,204,212,215,217,219,224,233,241,257,279,307,328,331,342,349,354,364,369,370,373,377,379,387,395,425,426,430,431,441,453,455,456,466,473,482,512,532–534 |

| | | | | |
|---|---|---|---|---|
| **107** | | | **Culturally acceptable / appropriate foods** | 19,44,46,47,52,57,64,67,68,82–84,86,96,102,114,132,148,165,174,179,191,200,205,212,215,217,219,224,233,245,254,271,283,322,328,329,331,347,369,385,387,391,434,444,447,454,464,467,482,490,495,501,511,512 |
| **108** | | | **Taste/sensory appeal of food** | 17,73,82,96,114,128–130,135,136,157,165,179,180,190,197,199,205,211,217,219,233,234,241,257,268,269,279,283,296,326,337,349,352,366,369,370,372–374,377,378,385,387,391,394,396,411,413,419,425,428,429,431,455,512,534,535 |
| **109** | | | **Food taboos** | 19,46,61,64,67,68,71,79,83,97,99,102,107,114,116,132,133,136,148,164,165,208,209,217,268,269,271,283,320,322,336,373,390,399,444,454,456,467,480,482,495,498,501,511,532,536,537 |
| **110** | | | **Attitudes towards food** | 17,50,62,82,85,87,102,103,105,114,129,157,179,180,191,200,202,204,217,224,266,279,296,302,329,337,345,347,362,373,377,385,388,394,454,463,466,473,487,505,529,531,538 |
| **111** | | | **Dietary preferences** (e.g., vegan, vegetarian) | 19,45,50,64,78,84,86,217,326,385,387,420,439,495,498,531 |
| **112** | | | **Acceptability of product (general)** | 194,211,328,372,377,397,398,400 |
| **113** | | | **Acceptability of product (fortified)** | 53,110,128–130,157,283,370,378,393,413,419,421,428,439,535,538 |
| **114** | | | **Perceptions about suitability of food based on gender** (e.g., some foods such as meat perceived as being important for men) | 47,49,50,64,78,82,102,114,136,200,241,498 |
| **115** | | | **"Prestige foods"** | 82,114,179,192,219,424 |
| **116** | | | **Perceptions about suitability of food based on age** (e.g., fruits perceived as children's food) | 87,114,136,331 |
| **117** | **(12) Social norms** | | **Socio-cultural food beliefs/expectations** (e.g. religious fasting, galactagogue feeding practices for lactating women, food aid stigma, taboos) | 17,19,44,46,50,52,57,61,64,67,68,71,74,75,79,82,83,85,93,95,99,102,106,107,114,128,129,133,136,144,148,164,179,193,208,209,212,215,217,224,263,268,269,322,327,329,354,366,369,373,385,391,399,431,444,447,454,456,460,467,468,490,495,498,501,505,511,529,532,537,539–546 |
| **118** | | | **Household/family food preferences** (e.g., spouse, children, parents, other household members) | 44,45,47,49–52,61,64,65,68,69,73–75,77,78,82,86,93–95,104,106,112,133,179,188,192,207,211,212,215,218,219,224,233,241,254,257,296,302,307,342,364,370,372,473 |
| **119** | | | **Peer influence** | 17,19,46,82,86,87,94,96,103,112,114,117,179,195,208,215,233,241,326,331,349,369,373,374,391,394,429,444,455,459 |
| **120** | | | **Socio-cultural food sharing expectations** (e.g., wontok, socio-cultural gatherings) | 19,21,44,47,54,57,82,86,93,102,114,117,122,180,194,215,249,290,339,393 |
| **121** | | | **"Pester power" (children's food preferences)** | 51,52,57,68,77,82,86,94,190,197,211,217,218,224,254,364,376,377 |
| **122** | | | **Household/family morbidities / health needs (e.g., family member with diabetes)** | 51,52,64,78,192 |
| **123** | **(13) Social capital** | **Social capital/-networks** | **Social networks/support between neighbours/family/friends,** including interdependent relationships for food procurement | 20,21,44,55,56,63–65,69–71,77,79,84,86,94,95,99,101,103,106,112,113,117,119,120,122,136,172,173,178,184,188,190,192–194,203,210,213,216,248,250,260,273,275,277,330,376,393,437,453,459,487,540,541,546–549 |
| **124** | | | **Food sharing/gifting of food** (including kin/community food sharing) | 19,44,54,57,64,69,70,75,86,93,94,99,102,113,114,120,122,138,146,202,205,210,214,215,248,268,272,290,330,339,374 |
| **125** | | | **Sharing of nutrition knowledge / how or where to access resources** | 54,57,67,70,71,83,86,91,95,110,112,133,192,193,202,208,212,269,273,331,339,373,391,444,454,459,529 |

| | | | | |
|---|---|---|---|---|
| **126** | | | **Rapport with food vendors** | [54,59,86,112,120,122,188,202,211,212,217,254,257,268,290,302,307] |
| **127** | | | **Level of rapport/trust between community members** | [20,54,88,97,112,136,260,268,273,307,487] |
| **128** | | | **Women's collaborative groups – formal** (e.g. farming group membership, self-help group, financial/savings/credit group) | [20,65,94–96,110,195,198,326] |
| **129** | | | **Women's collaborative groups – informal** (e.g., between neighbours, co-wives, female family members, allomaternal support, coorperative labour networks) | [20,69,112,188,190,205,210,248,376] |
| **130** | **(14) Convenience** | **Perceived time / effort required to…** | **Prepare food** | [20,45,50,52,57,82,86,88,97,100,103,129,130,148,179,188,190,192,197,202,206,207,211,212,214,215,217–219,224,241,245,257,266,298,301,302,328,329,342,366,372,377,396,430,431,473] |
| **131** | | | **Acquire food** | [20,88,91,92,99,135,138,166,190,197,202,205,212,214,217,218,224,249,254,257,302,369,376,381,542,550–552] |
| **132** | | | **Cook food** | [45,50,86,88,97,100,129,148,179,188,190,192,202,207,212,214,217,218,245,266,296,329,349,396] |
| **133** | | | **Produce / grow food** | [20,57,88,94,96,97,100,136,195,352] |
| **134** | | | **Consume food** | [17,47,93,129,130,148,214,329] |
| **135** | | | **Preserve food** | [195] |
| **136** | | **Time allocation** | **Time allocation for food acquisition / preparation / cooking** | [20,22,47,65,68,72,82,88,100,117,138,185,190,193,197,206,257,329,372,374,429] |
| **137** | | **Storage/-cooking facilities** | **Lack of storage, including cold storage** (e.g. refrigeration / cold storage, sealed containers) | [65,97,112,113,128,130,188,193,212,215,217] |
| **138** | | | **Limited / no cooking facilities or supplies** (e.g., no kitchen, firewood, gas, oil, clean water) | [47,70,113,211,212,215,257,310] |
| **139** | **(15) Food safety perspectives** | | **Concerns about chemical and microbiological contamination** (e.g., agrochemicals, pesticides, herbicides, adulteration, environmental contamination, bacterial etc.) | [52,54,57,75,82,113,116,199,211,212,217,233,241,254,257,282,301,302,307,342,373,379,391,399,400,551,552] |
| **140** | | | **Concerns about vendor sanitation** | [59,82,86,199,211,212,217,254,257,302,374,376,381] |
| **141** | | | **Safety concerns related to packaging and storage** | [59,82,128,202,211] |
| **142** | | | **Safety concerns about re-labelling beyond use-by-date** | [113,376] |
| **143** | | | **Safe food conflated with healthy foods** | [52,82,257] |
